# PERSISTENT IMMUNITY AFTER MILD SARS CoV-2 INFECTION - THE CoNAN-LONG TERM STUDY -

**DOI:** 10.1101/2022.07.05.22277237

**Authors:** Clara Schnizer, Nico Andreas, Wolfgang Vivas, Thomas Kamradt, Michael Baier, Michael Kiehntopf, Stefan Glöckner, André Scherag, Bettina Löffler, Steffi Kolanos, Joel Guerra, Mathias W. Pletz, Sebastian Weis, the CoNAN study group

## Abstract

**Objectives:** Understanding persistent cellular and humoral immune responses to SARS-CoV-2 will be of major importance to terminate the ongoing pandemic. Here we assessed long-term immunity in individuals with mild COVID-19 up to one year after a localized SARS-CoV-2 outbreak.

**Methods:** CoNAN was a longitudinal population-based cohort study performed 1.5 months, 6 months and 12 months after a SARS-CoV-2 outbreak in a rural German community. We performed a time series of five different IgG immunoassays assessing SARS-CoV-2 antibody responses on serum samples from individuals that had been tested positive after a SARS-CoV-2 outbreak as well as in control individuals who had a negative PCR result. These analyses were complemented with the determination of spike-antigen specific T_H_ cell responses in the same individuals.

**Results:** All infected participants presented as asymptomatic or mild cases. Participants initially tested positive for SARS-CoV-2 infection either with PCR, antibody testing, or both had a rapid initial decline in the serum antibody levels in all serological test but showed a persisting and robust T_H_ cell immunity as assessed by the detection of SARS-CoV-2 specificity of T_H_ cells for up to one year after infection.

**Conclusion:** Our data support the notion of a robust T cell immunity in mild and asymptomatic cases of SARS-CoV-2 up to one year after infection. We show that antibody titers decline over one year, but considering several test results, complete seroconversion is rare.

**Trial Registration:** German Clinical Trials Register DRKS00022416.

**Funding:** Sondervermögen „Corona” of the Thuringian Ministry for Economic Affairs, Science and Digital Society. SV-Kapitel 82 30 Titel 68205 # 5526/32-4-2.

## INTRODUCTION

Understanding immunity to SARS-CoV-2 will be of major importance to terminate the ongoing pandemic (1, 2). A growing body of evidence shows that SARS-CoV-2 infections lead to the induction of a broad humoral and cellular immune response that correlate with disease severity (1, 3, 4). This immune response is affected by individual host factors such as age, sex and comorbidities similar to other infectious disease (5–7).

After infection, seroconversion *i*.*e*. the development of antibodies against structural proteins of the virus such as spike-protein including the receptor binding domain (RBD) or the nucleocapsid protein of the virus has been demonstrated in 50 to 100% of patients. However, depending on the studied population its utility for the assessment of immunity has been questioned (3, 8–11). In contrast, neutralizing antibodies that are not measured routinely have been show do persist for up to one year (12, 13).

Rapidly after infection, also a T cell-mediated immunity is mounted that directly controls disease severity (3, 14, 15). Higher numbers of CD4^+^ and CD8^+^ T cells were associated with a milder course of disease (16, 17). In line with this, a higher degree of T cell activation with concomitant decreased numbers of T cells was correlated with an increased disease severity (17–19). Furthermore, COVID-19 severity was associated with a stronger inflammatory T cell-mediated cytokine response against S-, M- or N-proteins early after infection (20, 21). Additionally, infection by SARS-CoV-2 also provokes a specific memory T_H_ cell response that has shown to be stable at least for several months (15, 19, 22–24). Surprisingly, only few studies report follow-ups to one year after infection (25, 26). Notably, the huge majority of studies over a time period beyond six months follows hospitalized cases of Covid-19 (27) leading to an overrepresentation of medium or severe cases of COVID-19. Only a few studies report antibody or T cell responses after mild or even asymptomatic cases after more than six months after infection (14, 28, 29).

While disease severity correlates with levels of SASR-CoV-2-specific T cells and serum antibodies early after infection (30), in mild cases a stable T cell response appears to be preserved as well up to one year after infection (26, 31). It appears, that these cases are most important to understand the role of antibody and T-cell mediated herd immunity (32) and robust protection after vaccination (33). Due to the global vaccination campaign that started in 2021 and multiple SARS-CoV-2 infection waves, it becomes increasingly challenging to enroll and follow up infected subjects in the Western World without vaccination or re-infection which allows to assess the natural long-term course of infection. Thus, long-term data on the natural course of immunity after a single SARS-CoV-2 infection are scarce

The CoNAN study was a prospective longitudinal population-based study enrolling participants living in the small rural German community of Neustadt-am-Rennsteig, Germany starting in May 2020. After a local SARS-CoV-2 outbreak in the community and a 14-day quarantine of the entire village a field study was performed (8). This included sampling 1.5; 6 and 12 months after the outbreak. The combination of an isolated location and the well documented and controlled SARS-CoV-2 outbreak are unique features of this study allowing to assess the long-term immunity of SARS-CoV-2 infections without major biases. Here we report the long-term immunity in previously SARS-CoV-2 infected participants and healthy controls from the CoNAN study.

## RESULTS

### PARTICIPANT CHARACTERISTICS

The study flow chart of all three visits of the CoNAN study is shown in *Figure (Fig.) 1*. A total of 626 of the 883 community inhabitants (71%) participated in the first round of the study in April 2020. Of those, 162 individuals with a prior positive SARS-CoV-2 PCR and/or antibody titer (“infected”) and age and sex matched non-infected controls were invited via mail to participate in the 2^nd^ and 3^rd^ visit. Of the initial 626 participants, 146 individuals took part in the second visit in October 2020, and 224 in the third visit in April 2021. There were 132 individuals that participated in all three rounds. Antibody levels were determined in all of these participants. T cell analysis was performed in all previously infected participants as well as in randomly chosen previously non-infected individuals. The detailed characteristics of the subjects are given in *Table 1, 2* and *Supplementary (Suppl.) Table 1*.

**Table 1:** Characteristics of the analyzed seropositive participants with matched samples from all three time points.

| Characteristics | All Antibody positive, (n=40) | PCR-positive (n=14) | PCR negative (n=26) |
| --- | --- | --- | --- |
| Age [mean, SD] | 61,55 (12,05) | 66,93 (13,04) | 58,65 (10,64) |
| Median [Min, Max] | 62 (36, 83) | 68 (39, 83) | 57 (36, 76) |
| Male | 22, (55,0%) | 7, (50,0%) | 15, (57,69%) |
| Female | 18, (45,0%) | 7, (50,0%) | 11, (42,31%) |
| **Chronic Disease Category (no, (%))** |  |  |  |
| BMI (mean, SD) | 27,4 (4,43) | 27,61 (4,52) | 27,29 (4,47) |
| Arterial Hypertension | 20, (50,0%) | 6, (42,86%) | 14, (53,85%) |
| Myocardial Infarction | 3, (7,5%) | 2, (14,29%) | 1, (3,85%) |
| Congestive Heart Failure, CHD | 4, (1,0%) | 3, (21,43%) | 1, (3,85%) |
| pAVK | 1, (2,5%) | 0, (0%) | 1, (3,85%) |
| Stroke | 1, (2,5%) | 0, (0%) | 1, (3,85%) |
| Chronic Lung Disease | 3, (7,5%) | 2, (14,29%) | 1, (3,85%) |
| Autoimmune Disease/Immunodeficiency | 2, (5,0%) | 1, (7,14%) | 1, (3,85%) |
| Liver Disease | 0, (0%) | 0, (0%) | 0, (0%) |
| Diabetes Mellitus | 3, (7,5%) | 2, (14,29%) | 1, (3,85%) |
| Chronic Renal Disease | 4, (1,0%) | 2, (14,29%) | 2, (7,69%) |
| Tumor | 0, (0%) | 0, (0%) | 0, (0%) |
| Chronic Wounds, Eczema | 0, (0%) | 0, (0%) | 0, (0%) |
| Chronic Viral Infection | 0, (0%) | 0, (0%) | 0, (0%) |
| Other Disease | 7, (17,5%) | 1, (7,14%) | 6, (23,08%) |
| Smoker | 5, (12,5%) | 0, (0%) | 5, (19,23%) |
| Former Smoker | 3, (7,5%) | 1, (7,14%) | 2, (7,69%) |
| **Antibody tests** |  |  |  |
| CoNAN 1 (Median, IQR) | 6 (6, 6) | 6 (6, 6) | 6 (5, 6) |
| CoNAN 2 (Median, IQR) | 4 (3, 5) | 4,5 (3,75; 5) | 3 (2, 4) |

The CoNAN long-term study
| CoNAN 3 (Median, IQR) | 4 (3, 4) | 4 (4; 4,25) | 3,5 (3; 4,25) |
| --- | --- | --- | --- |
| Clinical Symptoms (outbreak) |  |  |  |
| Fever | 9, (22,5%) | 4, (28,57%) | 5, (19,23%) |
| Cough | 18, (45,0%) | 10, (71,43%) | 8, (30,77%) |
| Nose congestion | 7, (17,5%) | 3, (21,43%) | 4, (15,39%) |
| Dyspnoe | 7, (17,5%) | 4, (28,57%) | 3, (11,54%) |
| Fatigue | 18, (45,0%) | 9, (64,29%) | 9, (34,62%) |
| Joint Pain | 14, (35,0%) | 8, (57,14%) | 6, (23,08%) |
| Sweating/Chills | 10, (25,0%) | 5, (35,71%) | 5, (19,23%) |
| Taste disorder | 16, (40,0%) | 9, (64,29%) | 7, (26,92%) |
| Smell disorder | 9, (22,5%) | 6, (42,86%) | 3, (11,54%) |
| Diarrhea, Vomiting, Abdominal Pain | 6, (15,0%) | 4, (28,57%) | 2, (7,69%) |
| Admitted to Hospital for Covid | 8, (20,0%) | 6, (42,86%) | 2, (7,69%) |
| Admitted to Intensive Care Unit for Covid | 1, (2,5%) | 0, (0%) | 1, (3,85%) |

**Table 2:**
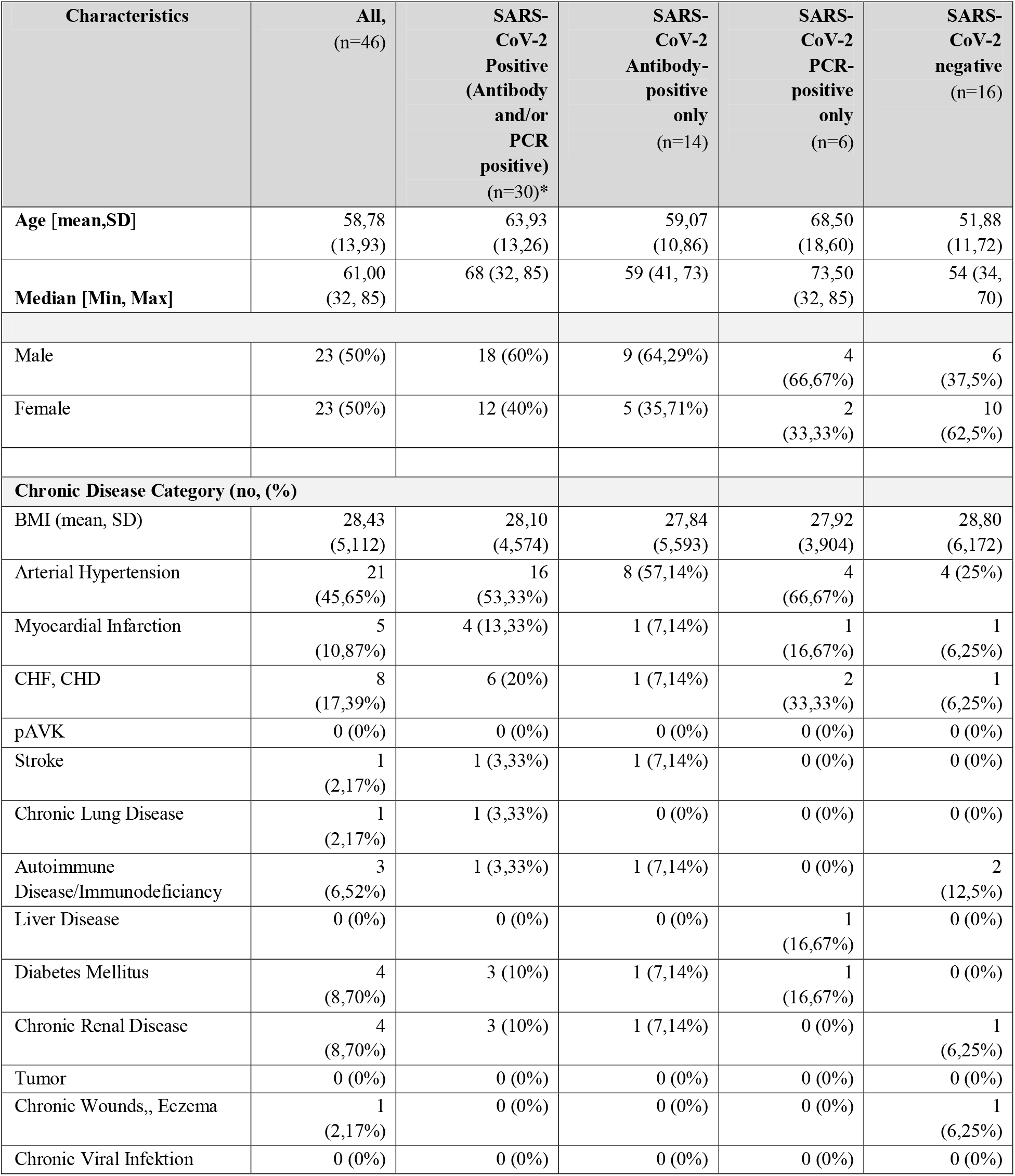
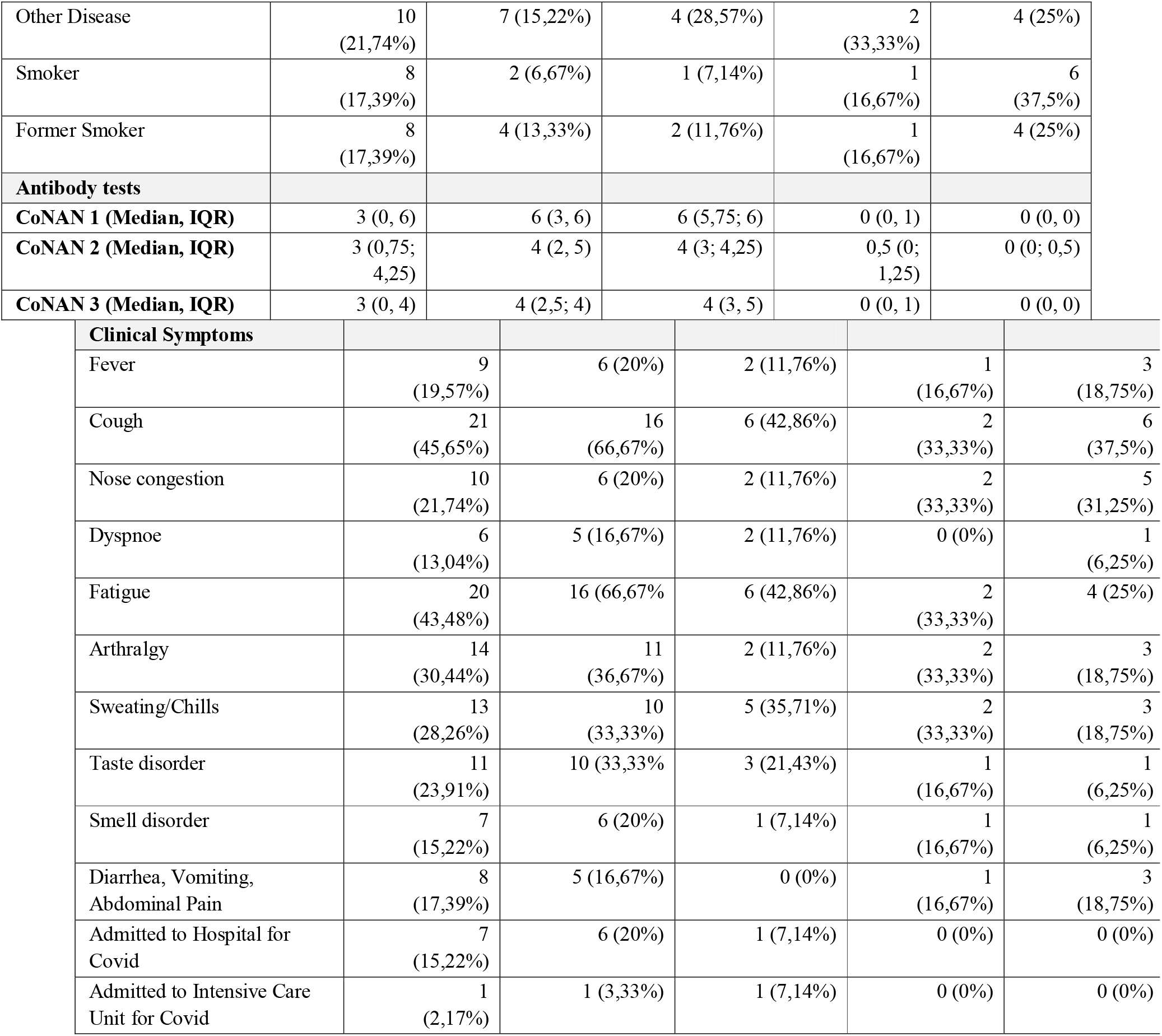
Participants characteristics of the T_H_-cell longitudinal cohorts. * 10 participants were PCR and antibody positive) <u>Abbreviations:</u> no..number; SD..standard deviation

| Characteristics | All,<br>(n=46) | SARS-CoV-2<br>Positive<br>(Antibody<br>and/or<br>PCR<br>positive)<br>(n=30)* | SARS-CoV-2<br>Antibody-positive<br>only<br>(n=14) | SARS-CoV-2<br>PCR-positive<br>only<br>(n=6) | SARS-CoV-2<br>negative<br>(n=16) |
| --- | --- | --- | --- | --- | --- |
| **Age [mean,SD]** | 58,78<br>(13,93) | 63,93<br>(13,26) | 59,07<br>(10,86) | 68,50<br>(18,60) | 51,88<br>(11,72) |
| **Median [Min, Max]** | 61,00<br>(32, 85) | 68 (32, 85) | 59 (41, 73) | 73,50<br>(32, 85) | 54 (34,<br>70) |
| Male | 23 (50%) | 18 (60%) | 9 (64,29%) | 4<br>(66,67%) | 6<br>(37,5%) |
| Female | 23 (50%) | 12 (40%) | 5 (35,71%) | 2<br>(33,33%) | 10<br>(62,5%) |
| **Chronic Disease Category (no, (%))** |  |  |  |  |  |
| BMI (mean, SD) | 28,43<br>(5,112) | 28,10<br>(4,574) | 27,84<br>(5,593) | 27,92<br>(3,904) | 28,80<br>(6,172) |
| Arterial Hypertension | 21<br>(45,65%) | 16<br>(53,33%) | 8 (57,14%) | 4<br>(66,67%) | 4 (25%) |
| Myocardial Infarction | 5<br>(10,87%) | 4 (13,33%) | 1 (7,14%) | 1<br>(16,67%) | 1<br>(6,25%) |
| CHF, CHD | 8<br>(17,39%) | 6 (20%) | 1 (7,14%) | 2<br>(33,33%) | 1<br>(6,25%) |
| pAVK | 0 (0%) | 0 (0%) | 0 (0%) | 0 (0%) | 0 (0%) |
| Stroke | 1<br>(2,17%) | 1 (3,33%) | 1 (7,14%) | 0 (0%) | 0 (0%) |
| Chronic Lung Disease | 1<br>(2,17%) | 1 (3,33%) | 0 (0%) | 0 (0%) | 0 (0%) |
| Autoimmune<br>Disease/Immunodeficiency | 3<br>(6,52%) | 1 (3,33%) | 1 (7,14%) | 0 (0%) | 2<br>(12,5%) |
| Liver Disease | 0 (0%) | 0 (0%) | 0 (0%) | 1<br>(16,67%) | 0 (0%) |
| Diabetes Mellitus | 4<br>(8,70%) | 3 (10%) | 1 (7,14%) | 1<br>(16,67%) | 0 (0%) |
| Chronic Renal Disease | 4<br>(8,70%) | 3 (10%) | 1 (7,14%) | 0 (0%) | 1<br>(6,25%) |
| Tumor | 0 (0%) | 0 (0%) | 0 (0%) | 0 (0%) | 0 (0%) |
| Chronic Wounds,, Eczema | 1<br>(2,17%) | 0 (0%) | 0 (0%) | 0 (0%) | 1<br>(6,25%) |
| Chronic Viral Infektion | 0 (0%) | 0 (0%) | 0 (0%) | 0 (0%) | 0 (0%) |
| Other Disease | 10<br>(21,74%) | 7 (15,22%) | 4 (28,57%) | 2<br>(33,33%) | 4 (25%) |
| Smoker | 8<br>(17,39%) | 2 (6,67%) | 1 (7,14%) | 1<br>(16,67%) | 6<br>(37,5%) |
| Former Smoker | 8<br>(17,39%) | 4 (13,33%) | 2 (11,76%) | 1<br>(16,67%) | 4 (25%) |
| **Antibody tests** |  |  |  |  |  |
| **CoNAN 1 (Median, IQR)** | 3 (0, 6) | 6 (3, 6) | 6 (5,75; 6) | 0 (0, 1) | 0 (0, 0) |
| **CoNAN 2 (Median, IQR)** | 3 (0,75; 4,25) | 4 (2, 5) | 4 (3; 4,25) | 0,5 (0; 1,25) | 0 (0; 0,5) |
| **CoNAN 3 (Median, IQR)** | 3 (0, 4) | 4 (2,5; 4) | 4 (3, 5) | 0 (0, 1) | 0 (0, 0) |
| **Clinical Symptoms** |  |  |  |  |  |
| Fever | 9<br>(19,57%) | 6 (20%) | 2 (11,76%) | 1<br>(16,67%) | 3<br>(18,75%) |
| Cough | 21<br>(45,65%) | 16<br>(66,67%) | 6 (42,86%) | 2<br>(33,33%) | 6<br>(37,5%) |
| Nose congestion | 10<br>(21,74%) | 6 (20%) | 2 (11,76%) | 2<br>(33,33%) | 5<br>(31,25%) |
| Dyspnoe | 6<br>(13,04%) | 5 (16,67%) | 2 (11,76%) | 0 (0%) | 1<br>(6,25%) |
| Fatigue | 20<br>(43,48%) | 16 (66,67%) | 6 (42,86%) | 2<br>(33,33%) | 4 (25%) |
| Arthralgy | 14<br>(30,44%) | 11<br>(36,67%) | 2 (11,76%) | 2<br>(33,33%) | 3<br>(18,75%) |
| Sweating/Chills | 13<br>(28,26%) | 10<br>(33,33%) | 5 (35,71%) | 2<br>(33,33%) | 3<br>(18,75%) |
| Taste disorder | 11<br>(23,91%) | 10 (33,33%) | 3 (21,43%) | 1<br>(16,67%) | 1<br>(6,25%) |
| Smell disorder | 7<br>(15,22%) | 6 (20%) | 1 (7,14%) | 1<br>(16,67%) | 1<br>(6,25%) |
| Diarrhea, Vomiting,<br>Abdominal Pain | 8<br>(17,39%) | 5 (16,67%) | 0 (0%) | 1<br>(16,67%) | 3<br>(18,75%) |
| Admitted to Hospital for<br>Covid | 7<br>(15,22%) | 6 (20%) | 1 (7,14%) | 0 (0%) | 0 (0%) |
| Admitted to Intensive Care<br>Unit for Covid | 1<br>(2,17%) | 1 (3,33%) | 1 (7,14%) | 0 (0%) | 0 (0%) |

For all three time points a matched analysis of antibodies of 40 participants was conducted. These were defined as being “seropositive” if at least two out of five performed serological tests. In a matched analysis of T_H_ cell immunity, we investigated 30 previously infected participants and 16 non-infected controls and excluded 10 participants that had been vaccinated or infected during the survey period. A comparison of the test performance between the five serological IgG assays in the participants is shown in *Suppl.Table 1*.

### LONGTERM ANTIBODY-RESPONSES TO SARS-CoV-2

In the population-wide CoNAN 1 study - in this manuscript referred to as 1.5-month time point-(8), 56 participants had been anti-SARS-CoV-2 antibody seropositive (AB+). Of these, 44 individuals participated in the 2^nd^ and 46 individuals in the 3^rd^ round of the study, respectively. Four participants had been vaccinated during the course of the study and were excluded. The remaining 40 participants were assessed for antibody course and are referred to as the “infected” group.

From the participants of the 3^rd^ visit, 161 had initially been tested negative (AB-) of which 40 (24,8%) participants became AB+. Of these, eighteen persons had been vaccinated against SARS-CoV-2. For one participant, the information on vaccination history was missing. Nine participants had a PCR-confirmed SARS-CoV-2 infection between the 2^nd^ and 3^rd^ visit. Further 21 participants had not been vaccinated against SARS-CoV-2.

The remaining 40 participants (median age 60.5 years (range 5-83, IQR 51,75-71), male n=24 (57,43%), female 18 (42,86 %)) were antibody positive in the first round and had no re-infection or vaccination against SARS-CoV-2. These were assessed in the longitudinal serology study. Participants characteristics are provided in *Table 2*.

In a first step, we assessed the course of the serum-antibody concentrations over one year after SARS-CoV-2 infection. Three out of five tests revealed discrete results normalized to a standard for all three time points (1. *EDI*, recognizing anti-nucleocapsid antibodies; 2. *Liason*, recognizing anti-spike antibodies and 3. *Maglumi Snibe*, recognizing anti-spike and anti-nucleocapsid antibodies), while one test (Euroimmune; recognizing anti-spike antibodies) provided an OD and semi-quantitative data and one test (Roche, recognizing anti-nucleocapsid antibodies) provided qualitative results only. As the missing standardization of the Roche test is a possible bias for tests performed at different time points, the course of the Roche test was not assessed further. During the one year observation period, the four quantitive tests showed a significant decline of the serum antibody concentrations (*Fig. 2A*). The extent varied between the individual tests and time points (shown in detail in *Fig. 2A & Suppl.Fig. 1A*). Others had shown that anti-nucleocapsid antibodies become undetectable as early as already eight months infection (34), suggestive of a shorter half-life of this antibody subset (35, 36). However, in our study the opposite was the case. When comparing the decline of antibody concentrations between the EDI (N) test and the Euroimmune (S) test between 1.5 and 6 months, the decrease in anti-nucleocapsid antibodies was less pronounced (mean_EDI_=0.84; SD_EDI_=0.37 *vs*. mean_EU_=0.21; SD_EU_=0.10; t-test p<0.001). This effect persisted after 12 months (mean_EDI_=0.31; SD_EDI_=0.11 *vs*. mean_EU_=0.21; SD_EU_=0.12; t-test p<0.001)(*Suppl.Fig. 2A*). For the early time-points, this was confirmed in the comparison of the EDI test with the Diasorin (S1/S2) test (mean_DS_=0.65; SD_EDI_=0.46; t-test p=0.05). Yet, after 12 months, the Diasorin assay had a less pronounced decline (mean_DS_=0.76; SD_EDI_=0.62; t-test p<0.001). This suggests a test-specific and not a antigen-specific effect. The data suggest a rapid waning of serum antibodies detected in some but not all tests during the first six months and a less rapid waning and preservation of antibodies within one year after SARS-CoV-2 infection.

**Figure 1:**
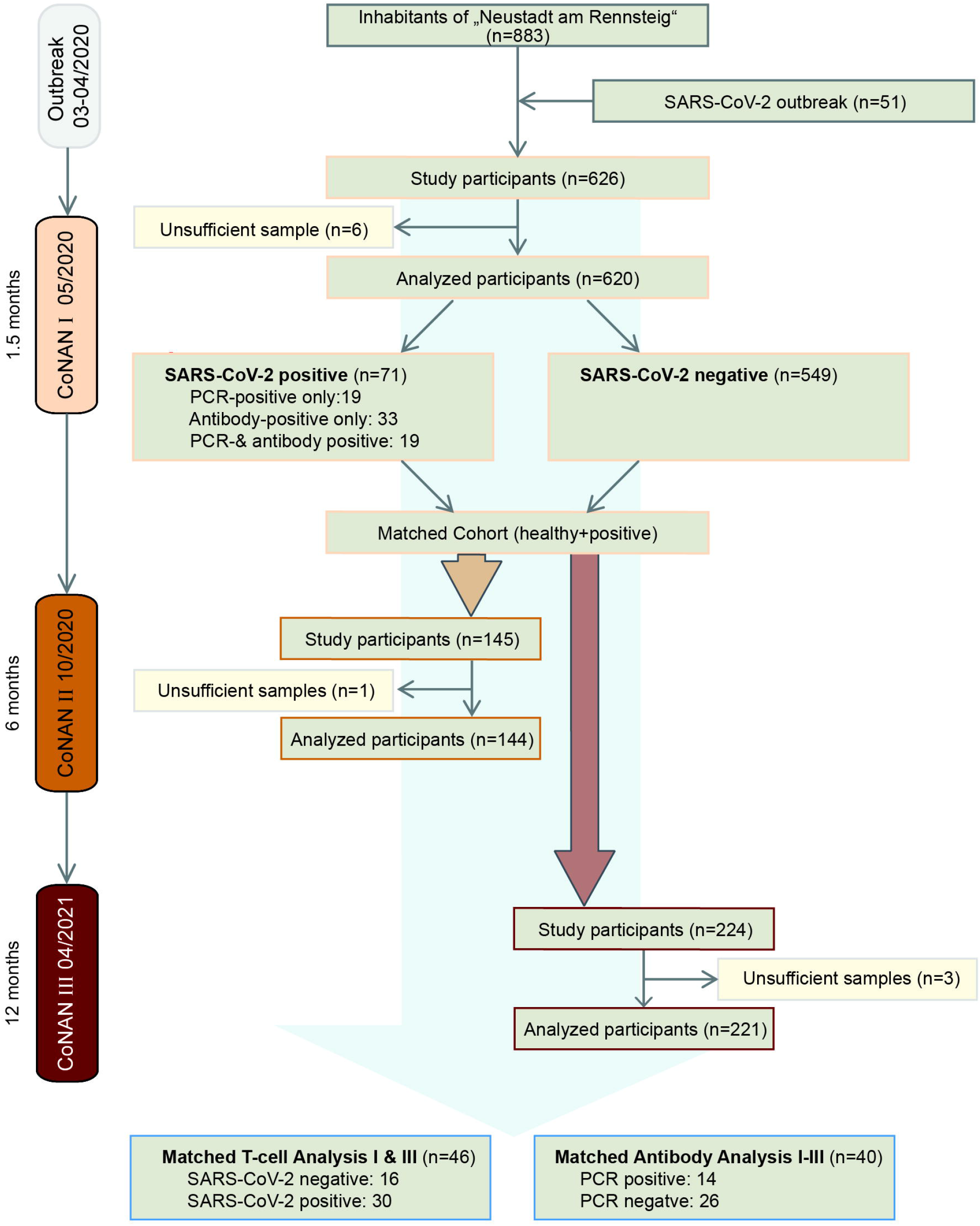
Flow chart of the CoNAN long-term study.

**Figure 2:**
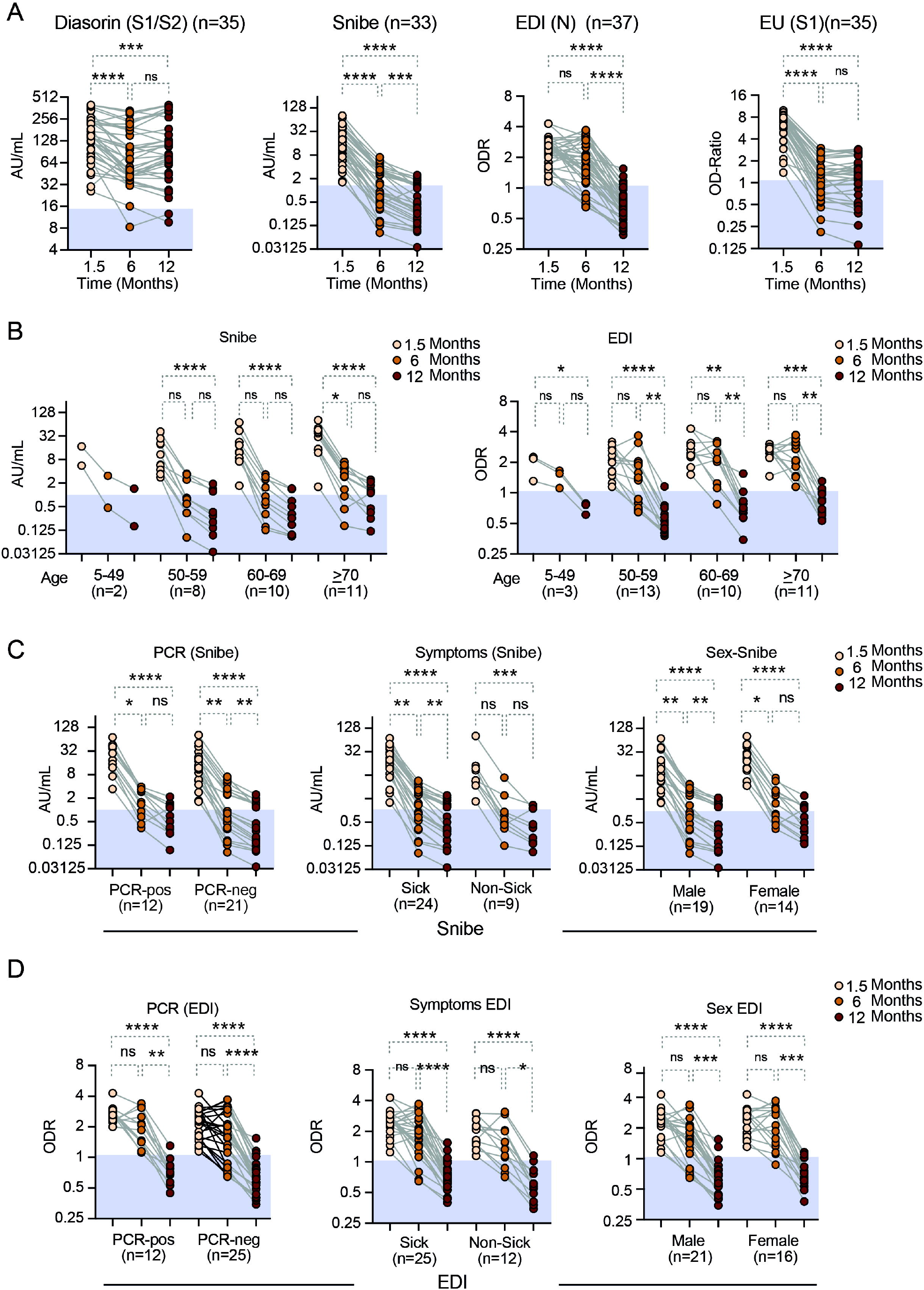
Anti-SARS-CoV-2 antibody levels over time as assessed with the three quantitative and one semiquantitative (EU) antibody tests as indicated **A**) all participants. Stratified by **B**) age for the Snibe (left panel) and EDI test (right panel). **C**) Results from the Diasorin Snibe antibody test stratified for PCR status (left panel), asymptomatic *vs*. symptomatic disease (middle panel) and sex (right panel). **D)** same as C for EDI test. N= number of individuals per group. Friedman test with Dunns post-hoc analysis. * p<0.05; ** p<0.01; *** p<0.001; **** p<0.0001. Abbreviations: AU.. arbitrary units, ns.. non-significant (p>0.05), Snibe: SN.. 2019-nCoV IgG kit (Snibe Co., Ltd., Shenzhen, China); ED.. EDI Novel Coronavirus SARS-CoV-2 IgG ELISA kit (Epitope Diagnostics Inc., San Diego, USA).

In a subgroup analysis, we also stratified the serum antibody concentrations with respect to age, PCR-positive *vs*. PCR-negative participants, the presence or absence of symptoms or the sex of the individuals. For all groups, the time course results remained unaffected by this stratification (*Fig. 2B,C,D; Suppl.Fig. 1B,C*). Notably, except three, all of the participants that initially had been tested seropositive-defined as at least two different positive assays-remained seropositive after one year.

### LONGTERM T CELL IMMUNITY TO SARS-CoV-2

To assess SARS-CoV-2 T cell-mediated immunity, we then analyzed S-protein-specific CD154^+^4-1BB^+^ cells among peripheral blood CD3^+^CD4^+^ T_H_ cells. This T cell-mediated immunity was determined in a matched cohort of 46 study individuals that participated in the first and third round of the study (*Fig. 1*). Of these, 16 individuals were non-infected and considered the control cohort. Thirty individuals were the infected cohort of which six were PCR-positive only, 10 were PCR-positive and antibody-positive after 1.5 months and 14 were antibody-positive only.

Characteristics of the participants in the longitudinal T cell study are provided in *Table 3*. To identify the spike-reactive T_H_ cells, we compared T_H_ cells restimulated with mixes of peptides covering the N-terminal part (*S.Pep1 (N)*) or the C-terminal part (*S.Pep2 (C)*) of the SARS-CoV-2 spike protein to T_H_ cells responding in presence of DMSO alone (*Fig. 3*). As shown in *Figure 3A & B* we could detect the presence of spike-specific CD154^+^4-1BB^+^ T_H_ cells among all CD4^+^CD3^+^ T_H_ cells at 1.5 months and still 12 months after infection (*Fig. 3A,B*). Notably, there was a slight, but significant reduction in the frequency of spike-specific T_H_ cells in the previously infected cohort over time (*Fig. 3C,D*). In only two participants (6.7%) the spike-specific T_H_ cell response had vanished after one year (*Fig. 3C, D*). When compared to SARS-CoV-2 negative participants, previously SARS-CoV-2 infected participants clearly showed an overall higher frequency of SARS-CoV-2 specific T cells at all time points that were investigated (*Fig. 3E, F*). In addition, when assessing the subgroup of antibody-positive (and PCR-negative) participants only, this trend persisted (*Fig. 3G, H*). Despite a persistent presence of spike-reactive T_H_ cells 12 months after infection in this group, a slight decrease in the frequency of these antigen-specific T_H_ cells among all T cells was also detectable (*Fig. 3H*). Interestingly, a significantly higher frequency of spike-specific T_H_ cells was detected in this antibody-only group, when compared to individuals with an initially positive PCR-status without measurable antibody titers (*Suppl.Fig.3*). Notably, several healthy subjects, showed a T_H_ cell response against SARS-CoV-2 already at the beginning of the study which was maintained over time.

**Figure 3:**
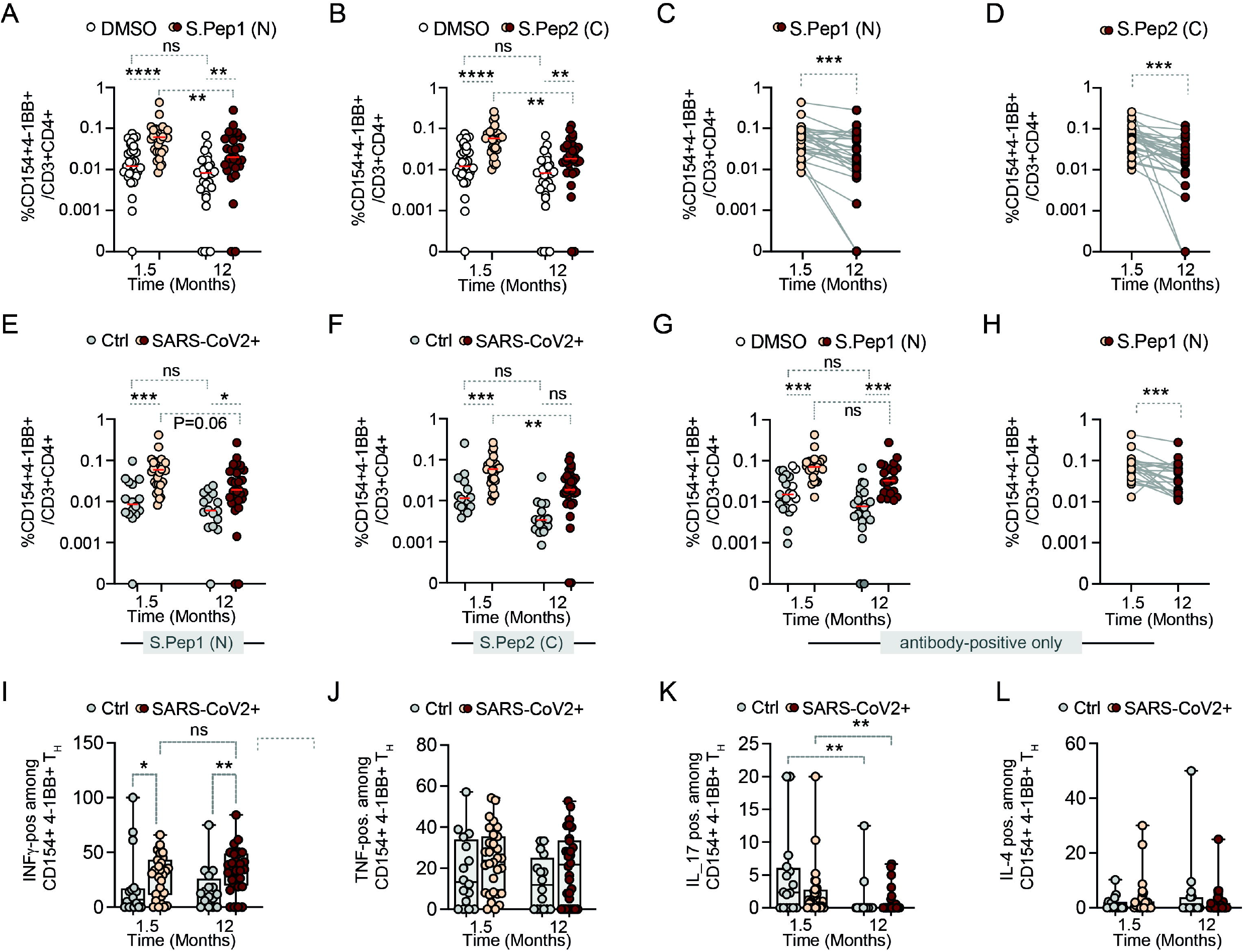
Longitudinal SARS-CoV-2 specific T_H_ cell response. **A**) % CD154^+^4-1BB^+^ T_H_ cells from previously infected participants stimulated with DMSO or S.Pep.1 (N-terminal). **B**) % of 154+4-1BB+ T_H_ cells stimulated with DMSO or S.Pep.2 (C-terminal). **C and D**) Time course of SARS-CoV-2 specific T_H_ cells stimulated with **C**) S.Pep.1 (N) and **D**) S.Pep.2 (C). **E and F**) % CD154^+^4-1BB^+^ T_H_ cells stimulated with **E**) S.Pep.1 (N) and **F**) S.Pep.2 (C) in SARS-CoV-2 negative (Ctrl) *vs*. positive (SARS-CoV-2+) participants. **G** and **H**) % CD154^+^4-1BB^+^ T_H_ cells in a subset of antibody positive participants only. **I-L**) Positive CD154^+^4-1BB^+^ T_H_ after stimulation with S.Pep.1 for the cytokines **I**) INFγ, **J**) TNF, **K**) IL-17 and **L**) IL-4. Dots represent individual participants. Time indicates months after the SARS-CoV-2 outbreak. Wilcoxon-matched-pairs test for two matched groups and Kruskal-Wallis with Dunns post-hoc for other analysis with more than two groups.). * p<0.05; ** p<0.01; *** p<0.001; **** p<0.0001. <u>Abbreviations:</u> AB.. antibody; Ctrl.. controls (non-infected participants); DMSO.. dimethyl sulfoxide; IL.. Interleukin; INF.. Interferon; ns.. non-significant (p≥0.05).

In conclusion, individuals with initially detectable antibody levels showed a higher T_H_ cell response after 12 months than individuals with an antibody titer below detection threshold despite of a PCR-confirmed infection. When we assessed intracellular cytokine expression of INFγ, TNF, IL-4 and IL-17A (*Fig. 3I-L; Suppl.Fig. 3*), we observed that significantly elevated initial levels of INFγ expressing SARS-CoV-2 specific T_H_ cells in inpreviously infected patients remained elevated up to one year after infection (*Fig. 3I; Suppl.Fig. 3*), while the expression of TNF, IL-4 or IL-17A was not increased at any time point (*Fig. 3J-L; Suppl.Fig. 3*). Overall, despite the slight decrease in particular subgroups, the data suggest that a T_H_ cell-mediated immunity after SARS-CoV-2 infection prevails for at least one year and contains a robust and maintained specific T_H1_ cell immunity.

To gain a further unbiased perspective on the immunity toward the spike proteins as represented by the S.Pep.1 (N) or S.Pep.2 (C)-specific T_H_ response, we performed multi-dimensional flow cytometry analyses. Representative Uniform Manifold Approximation and Projection (UMAP) maps were color-coded according to the resulting clusters using the FlowSOM algorithm (37) (*Fig. 4, Suppl. Fig. 4*). With the limitation that only small populations could be analyzed, the global high-dimensional analyses revealed diverse T cell activation status of SARS-CoV2 infected individuals in time (*Fig. 4A, B*). For both S1(N) and S2(C), we observed populations that resembles different phenotypes: *i)* resting cells (cluster 1: negative for activation markers and cytokines), *ii)* 4-1BB^+^ activated CD4^+^ T cells (cluster 2: 4-1BB^+^IFNγ^+^TNF_low_), and *iii)* CD154 4-1BB activated CD4 T cells (cluster 3 for S.Pep1 and cluster 4 for S.Pep2: CD154^+^4-1BB^+^IFN_γ_^high^TNF^low^). Moreover, a S2(C)-specific population depicting a CD154^low^4-1BB^high^ phenotype was observed (CD154^low^4-1BB^high^IFN ^high^TNF^high^) (*Fig. 4B*). For both S1(N) and S2(C) we only observed a marginal contribution of IL-4 and IL-17A. Despite obtaining these clusters, we did not detect any significant differences in infected individuals between 1.5 months and 12 months. This unbiased result supports our notion that the T_H_-specific response is maintained over time after SARS-CoV-2 infection.

**Figure 4:**
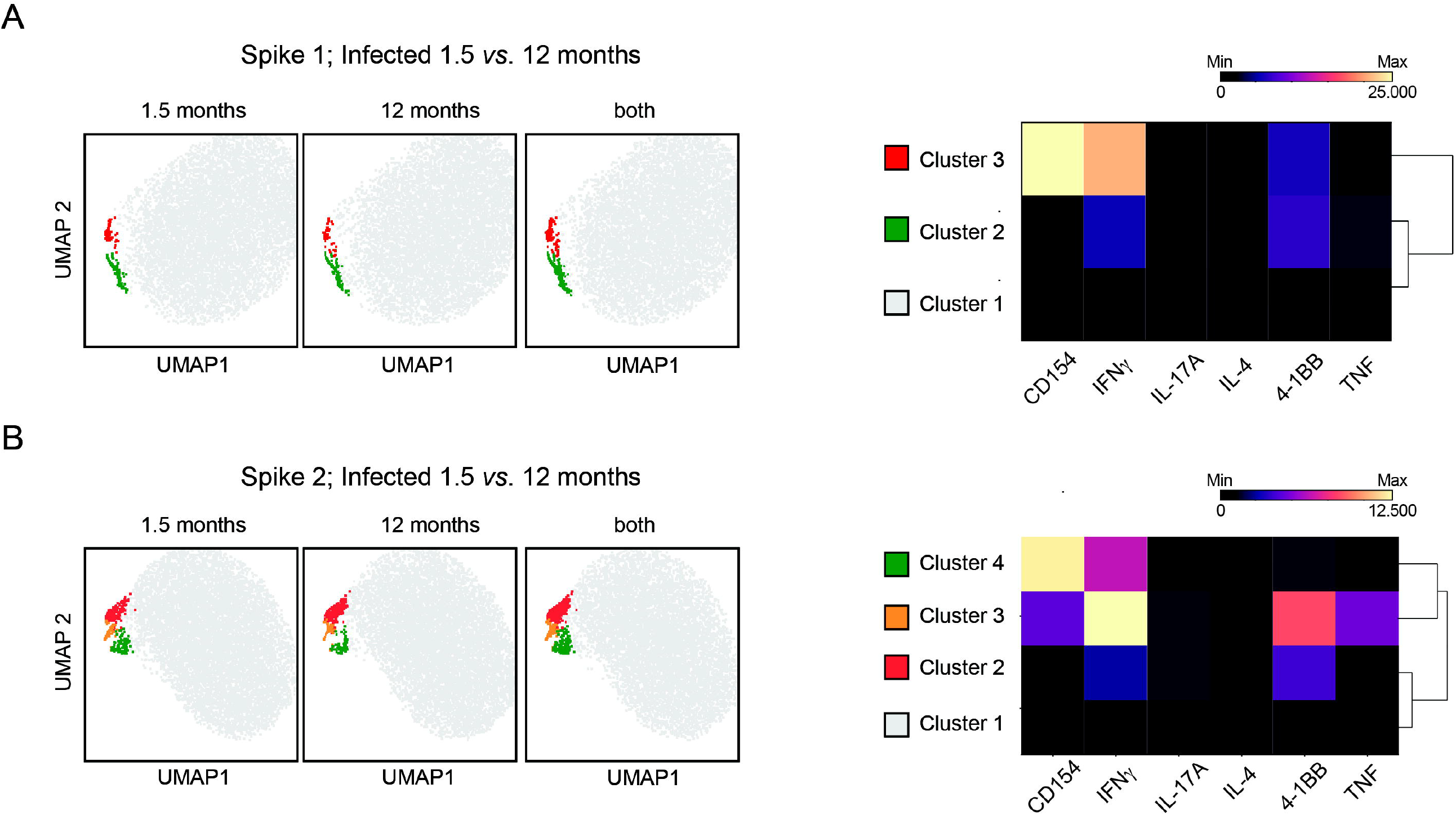
Multi-dimensional flow cytometry analyses using UMAP and FlowSOM clustering. Dot plots depict global UMAP projection pooled from CD3^+^CD4^+^ T cells from the study participants. For the first two maps, dot plots from each group are shown followed by a third dot plot with clusters identified with FlowSOM clustering using pooled CD3^+^CD4^+^ T cells from the compared groups. Heatmaps depict Median Fluorescence Intensity (MFI) values as indicated by clusters and markers. **A**) S.1 and **B**) S.2-specific T_H_ response in infected individuals for 1.5 *vs*. 12 months. <u>Abbreviations:</u> CD.. cluster of differentiation; IL.. interleukin; INF.. interferon; TNF.. tumor necrosis factor; UMAP..Uniform Manifold Approximation and Projection.

### T- AND B-CELL IMMUNITY TO SARS-CoV-2 CORRELATE-BUT ONLY WEAK

We then asked whether the 12-month antibody levels could be predicted by the T cell immune response mounted after 1.5 months and *vice versa*. Therefore, we correlated antibody concentrations and T cell immunity at 1.5 months and 12 months after infection. We assessed the Spearman correlation for the quantitative Diasorin, the Snibe, the EDI as well as for the semiquantitive Euroimmune assays with the percentage of the S1(N)-specific CD154^+^4-1BB^+^ T_H_ cells (*Fig. 5A-D*) and the S2(C)-specific CD154^+^4-1BB^+^ T_H_ cells (*Suppl.Fig. 5A-D*). None of the comparisons revealed a strong correlation. There was a moderate correlation (Spearman r=0.6-0.8) of the serological tests at 1.5 months to T_H_ cell responses at 1.5 months and 12 months after infection, but not at later time points (*Fig. 5A-D; Supp.Table 3*). When including all participants with T cell data regardless of the antibody status, this prevailed for the 1.5 months correlation. However, S2(C)-specific T_H_ cells moderately correlated with antibody concentrations for the EDI test at 1.5. months (r_EDI_=0.61; p<0.0001) (*Suppl.Fig. 5A*) and the IgG index of the Euroimmune assay (r_EU_=0.62; p<0.0001) (*Suppl.Fig. 5D*). The correlation to S1(N)-specific T_H_ cells was only weak (r_EDI_=0.39, p=0.05; r_EU_=0.48, p=0.0079 (*Fig. 5A, D*).

**Figure 5:**
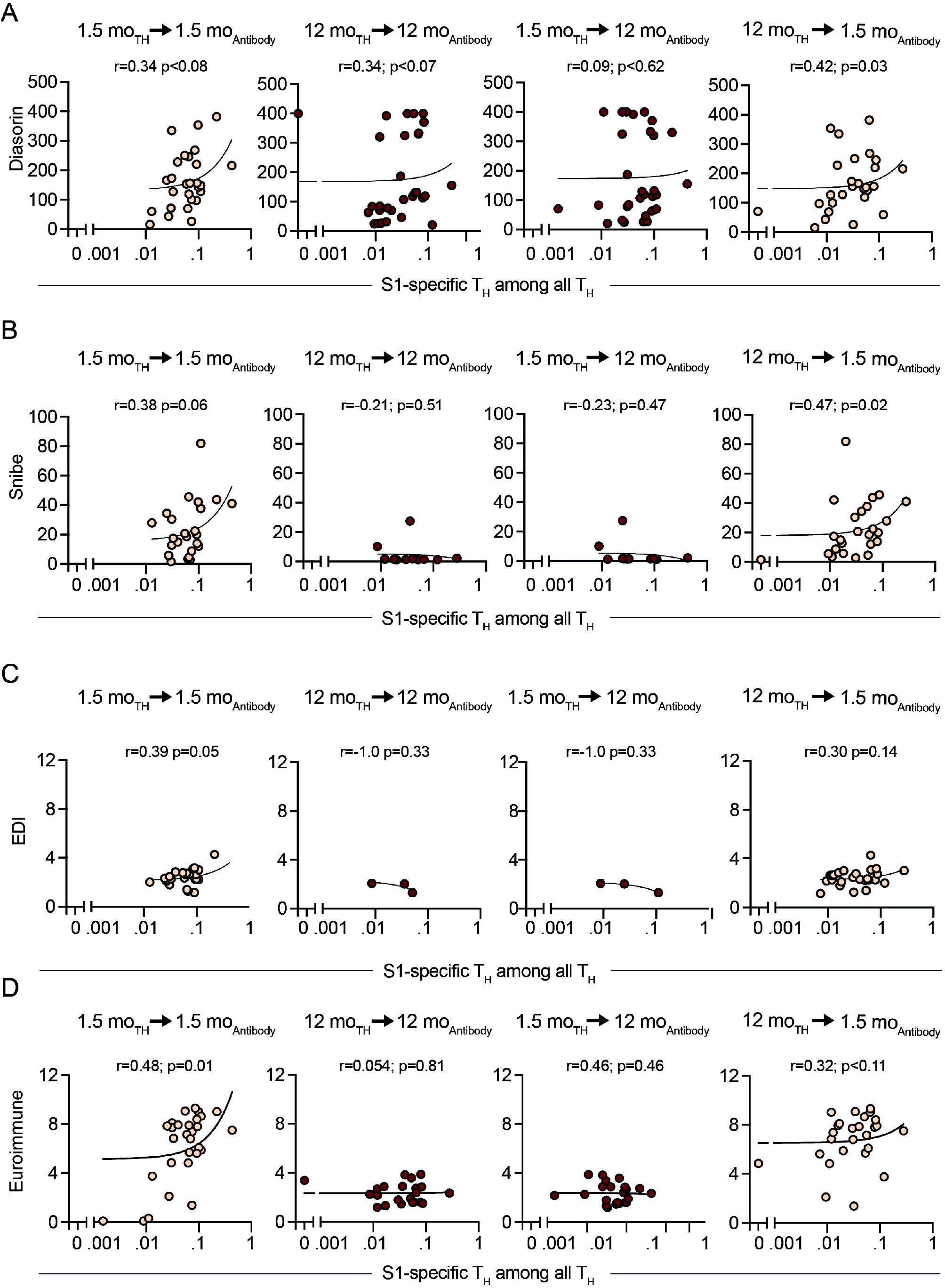
Spearman Correlation analysis of SARS-CoV-2 specific T_H_ cells (CD3^+^CD4^+^CD154^+^4-1BB^+^) after S.Pep-1 (N) stimulation with initially (1.5 months) positive serological test only. Comparisons were the different combinations of the 1.5-month and 12-month time points after the SARS-CoV-2 outbreak in Neustadt-am-Rennsteig. **A**) Liaison SARS-CoV-2 S1/S2 IgG CLIA DiaSorin, **B**) Maglumi 2019-nCoV IgG CLIA *Snibe*, **C**) EDI Novel Coronavirus COVID-19 IgG ELISA and **D**) Euroimmun Anti-SARS-CoV-2-ELISA (IgG). <u>Abbreviations:</u> mo.. month; T_H_.. T helper cells (CD3^+^CD4^+^). Dots indicate individual participants. Orange.. 1.5-month antibody data, Dark red.. 12-month antibody data.

## DISCUSSION

The *COVID-19 Outbreak in Neustadt-am-Rennsteig* (CoNAN) study was a longitudinal cohort study after a localized SARS-CoV-2 outbreak in a rural community in the federal state of Thuringia, Germany. We followed previously infected patients with predominant mild disease and uninfected participants for one year after the outbreak. We provide evidence for a persistent T-cell immunity and a prevailing antibody response over a one-year period. While the level of serum antibodies declined in a relevant manner during the first six months after infection, this decrease was slower during the subsequent six months. More importantly, seroconversion did not disappear completely and thereby, suggested a plateau of baseline antibody levels, albeit at low concentrations.

To date it is unclear, what determines protection against SARS-CoV-2 reinfection and which individuals are more likely to be persistently protected. In our initial study, only 50% of the previously infected individuals became seropositive (8). This assessment was performed approximately 1.5 months after infection and should represent the time period around the peak of an antibody response. Furthermore, it provides a good estimate about the humoral immune response directly after infection (35). Here, we show that serum antibody concentrations declined significantly over one year and thereby, support previous data (29). Of note, combining the results of all five antibody tests, which included the non-quantitative Roche assay, a seroconversion to negative was only observed in three participants. All others remained seropositive. Antibody decay appeared not to be linear with a more rapid decline directly after infection and a subsequent less pronounced waning as previously shown (38, 39). Modeling of humoral immune response suggest that antibody-mediated protection could be maintained for several years post infection even after mild disease (35, 39), which is supported by our data. Also, neutralizing capacity of the serum antibodies seemed to be preserved to a certain extent (12, 23, 40–43). Our data suggest that seropositivity of SARS-CoV-2 antibodies could be of shorter duration when compared to other severe corona virus-mediated disease. Several authors reported cellular immune response in patients after infection with the Severe Acute Respiratory Syndrome Corona Virus (SARS-CoV) or the Middle East Respiratory Syndrome (MERS)-CoV during previous epidemics (44). Both diseases are associated with a much higher mortality than SARS-CoV-2 (1). Long-term immunity against the related pathogens has been suggested on the basis of detectable serum antibodies up to two years after MERS infection (45, 46). Herein, memory T cell responses even persisted for several years (44, 47). Compared to MERS, SARS-CoV is associated with a longer memory response of specific T_H_ cells in two third of SARS survivors up to six years (48) and persisting neutralizing antibodies up to 17 years after infection (44, 49). With our study we contribute to the understanding of immune memory development in consequence of the related SARS-CoV-2 infection in mild and asymptomatic cases.

In this manuscript, we also assess the specific T_H_ cell memory response to SARS-CoV-2. SARS-CoV-2-specific memory T cells have been proposed to confer long term protection against SARS-CoV-2 re-infections (50). Our findings expand the observation of previous data obtained six months after mild infection that showed a persistent T-cell response against SARS-CoV-2 together with decreasing concentrations of spike- and nucleocapsid-specific antibodies (51). In general, strength and duration of an anti-SARS-CoV-2 T cell response depends on the severity of COVID-19 (52). Interestingly, the T cell response is directed against various epitopes (53, 54). The spike protein, a major cell entrance mediator of SARS-CoV-2 via ACE2 has been widely studied and used as targets for vaccination strategies, despite efficient responses were also induced against membrane and nucleocapsid proteins (55). While anti-nucleocapsid responses are dominated by CD8^+^ cytotoxic T cells (23), the anti-spike responses evoked are mainly mediated by T_H_ cells leading to follicular T helper cell circulation and antibody-producing B-cell responses (55–57). However, others have shown that SARS-CoV-2 T_H_ cell responses start decreasing as early as after six months and then persist at a lower magnitude (23, 55). Independent from the initial disease severity, the T_H_ cell-memory responses can mount protective T-cell responses mediated by IFNγ at any investigated later time point (58). In our study, T-cell responses were stable during the observation period. Surprisingly, especially in antibody-positive individuals that had no initial PCR confirmation of an infection, we observed a stable T_H_ cell response over one year. We can also show that there is an association between the early T-cell response and the late antibody levels. However, the strength of this association was only weak. This is in accordance with data obtained at 6 months after infection (23).

Additionally, SARS-CoV-2 specific T_H_ cells were detected in non-infected control participants. This phenomenon has already been reported and suggested to represent T_H_ cells that are cross-reactive to seasonal human coronaviruses (HCoVs) (24, 59, 60). Relative to this, SARS-CoV-2-specific T_H_ cells were increased after SARS-CoV-2 infection when compared to cross-reactive T_H_ cells (61). Whether this can be explained by potentially different periods that have passed since infection and thereby, indicates a waning T_H_ cell response or a qualitative difference, is currently unknown. Interestingly, the presence of these cross-reactive T_H_ cells has been shown to enhance SARS-CoV-2 immunity and improve the vaccination response (59). It is speculative, whether such a trend could explain the increased antibody titers in the elderly of our data set, which during a lifetime likely have encountered several HCoVs and might have built a lasting T-cell memory boosting antibody production (62, 63). Early studies about experimental infections of human volunteers with coronaviruses already showed that virus-induced antibody concentrations in the blood had been still increased after one year (64). While not completely preventing re-infection of such volunteers, the remaining immunity decreased the severity of the induced secondary infection (64). In how far the observed remaining antibody and T-cell responses correlate to a protection against re-infection or less disease severity in case of reinfection in SARS-CoV-2, however, remains speculative.

## CONCLUSIONS

Our data indicate the persistence of a T_H_ cell immunity even after mild SARS-CoV-2 and asymptomatic infection, which is a better predictor of long-term immune memory-mediated protection than initially measurable antibody titers. While antibody responses potentially wane below a detection minimum beyond one year after infection, specific T_H_ cell responses remain at a detectable level.

## METHODS

### Study design and enrollment

The CoNAN study (*COVID-19 Outbreak in Neustadt-am-Rennsteig*) was a prospective longitudinal population-based cohort study in Neustadt am Rennsteig a village in the Ilm-district in central Thuringia, Germany with 883 inhabitants in which a SARS-CoV-2 outbreak had occurred in Spring 2020. Due to the isolated location of the village, the extensive testing of the population, and the clear and controlled outbreak, Neustadt am Rennsteig is well suited to study the seroprevalence and potential development of immunity of SARS-CoV-2 infections. On March 22^nd^, 11 confirmed COVID 19 cases had been diagnosed in the district of which 6 (55%) were Neustadt residents with further 69 residents classified as contact persons. As a consequence, local public health authorities declared a 14-day quarantine on the entire village. With support of the local family physician, an outbreak containment team of the public health department conducted a mandatory mass screening using nasopharyngeal swabs starting on April 2^nd^ in which 865 SARS-CoV-2 PCR tests were performed. Health authorities reported 51 SARS-CoV-2 infections and three SARS-CoV-2-associated deaths in the community during the outbreak. All persons with positive PCR-results were defined as COVID-19 cases. The initiated containment measured controlled the outbreak the spread to neighboring villages was prevented. Quarantine on the village was lifted on April 5, 2020. For the CoNAN study, samples were taken at three defined time-points. The first sampling was performed from May 13 to May 16, 2020. A total of 626 participants were included. The results of the antibody testing have been published (8). The second sampling was performed from October 7 to October 9, 2020 and included the participants of the first round who had shown antibodies in at least two different IgG antibody assays and a control group matched after sex, age and co-morbidities, 145 participants in total. The third sampling was performed from April 13 to April 15, 2021 with the participants of round two along with some new participants, 224 in total (*Fig.1*). Participation in the study was voluntary and could be withdrawn at any time. Refusal to participate had no consequences. Participants were enrolled at a central study site that was set-up in the villages’ town hall or in rare cases if requested by home visits. After informed consent, questionnaires and blood samples were directly taken at the study site. All inhabitants of the community of Neustadt am Rennsteig regardless of age, gender or infection status were eligible for participation in the first phase. Individuals that do not reside in Neustadt am Rennsteig or that live in the adjacent community of Kahlert were not eligible for inclusion. Informed consent was provided by the participants or the parents/legal representatives. In the second and third phase of the study participants that had a proven infection with SARS either by SARS-CoV-2 PCR or antibody positivity in the first phase were invited along with an age-, sex and comorbidity matched control group. Inhabitants of the village that were not invited however could also perform antibody testing.

### Questionnaire

Participants completed a pseudonymized questionnaire directly at the study site during all three rounds. After re-assessing the original case report forms on paper, obvious errors were corrected, and duplicated entries were deleted. Plausibility checks of demographic data were performed. Symptoms were noted if reported. Strength and duration of symptoms was not weighted in the analysis of this manuscript. Self-reported information on a positive SARS-CoV-2 PCR test at the time point of the outbreak/quarantine initiation was double-checked with the information by the health department of the Ilm-district if the participants gave their permission on the consent form.

### SARS-CoV-2 antibody testing

Five serological tests were performed in all three rounds. Characteristics of the tests are provided in *Suppl. Table 1*. Two of the tests detected antibodies recognizing the S-antigen, one recognized the 2019-nCoV recombinant antigen and two tests recognized the N-antigen of SARS-CoV-2. Detection of SARS-CoV-2 IgG antibodies was performed with five different quantification methods, of which two were enzyme-linked immunosorbent assays (ELISA) and three were chemiluminescence-based immunoassays (CLIA/CMIA). All tests were carried out according to manufacturers’ instructions. For detailed information on assay characteristics and instruments used see *Suppl. Table 1*. Sensitivities and specificities are shown as provided by the manufacturer. The following assays were used; EDI Novel Coronavirus SARS-CoV-2 IgG ELISA kit (Epitope Diagnostics Inc., San Diego, USA), SARS-CoV-2 IgG ELISA kit (Euroimmun, Lübeck, Germany), SARS-CoV-2 S1/S2 IgG CLIA kit (DiaSorin, Saluggia, Italy), 2019-nCoV IgG kit (Snibe Co., Ltd., Shenzhen, China) and Elecsys Anti-SARS-CoV-2 kit (Roche, Basel Switzerland).

### Flow Cytometry Analysis

Peripheral Blood Mononuclear Cells (PBMCs) from 56 inhabitants of the village Neustadt am Rennsteig were analyzed for S-Protein-specific T_H_ cell response. DMSO as solvent of S-peptide mixes was used as control. S-peptide mixes 1 and 2 represent the S-Protein N-terminal part and C-terminal part, respectively. General reactivity was controlled by SEB/TSST1 stimulation. There were no non-responders. PBMCs were isolated by gradient density centrifugation on Biocoll solution (Bio&SELL GmbH, Germany) at 800 x g at room temperature (RT) for 20 min without brakes. PBMCs were washed with PBS and subsequently cryoconserved in medium containing Penicillin/Streptomycin (Sigma-Aldrich), 10 % DMSO (Sigma-Aldrich) and 50 % FCS (Sigma-Aldrich). For analysis, PBMCs were thawed and immediately washed with cell culture medium (supplemented with 10% human AB serum (PAN Biotech, Germany), penicillin/streptomycin). Upon recovery at 37°C for 2h, a maximum number of 5 × 10^6^ PBMCs were restimulated in cell culture medium containing 1 μg/mL recombinant anti-human CD28 antibody (clone CD28.2, BioLegend, RRID:AB_314303) and either 0.2 % DMSO (negative control), SARS-CoV-2 Spike glycoprotein PepMix 1 (S1, N-terminal coverage) or 2 (S2, C-terminal coverage) (both jpt, Germany). As high controls 10^6^ PBMCs were restimulated with 1 μg/mL TSST1 and 1 μg/mL SEB (both Sigma-Aldrich) in presence of 1 μg/mL recombinant anti-human CD28, or with anti-human CD3/CD28 beads (Gibco/Thermo Fisher Scientific, Lithuania) at a ratio of 1 bead/PBMC. After stimulation for 2 hrs, Brefeldin A (BioLegend) was added for another 14 hrs of incubation. Cells were shortly incubated with 1 mg/mL beriglobin followed by staining with anti-human CD3 Pacific Blue (clone UCHT1, BioLegend, RRID:AB_1595437) and anti-human CD4 Brilliant Violet 605 (clone RPA-T4, BioLegend RRID:AB_2564391). After 5 min at 4°C in the dark, Zombie Aqua fixable dead cells stain (BioLegend) was added, samples were mixed and incubated for another 10 min at 4°C in the dark. Incubation was stopped with PBA/E and the cells were fixed in 2% Formaldehyde/PBS at RT for 20 min, blocked with 1 mg/mL beriglobin/0.5% Saponin and intracellularly stained with anti-human CD154 APC (clone 24-31, BioLegend, RRID:AB_314832), anti-human CD137 PE/Cy7 (clone 4B4-1, BioLegend, RRID:AB_2207741), anti-human IFN_γ_ APC/Cy7 (clone 4S.B3, BioLegend, RRID: AB_10663412), anti-human TNF_α_ PerCP/Cy5.5 (clone MAb11, BioLegend, RRID: AB_2204081), anti-human IL-4 PE (clone MP4-25D2, Biolegend, RRID: AB_315129), and anti-human IL-17A FITC (clone BL168, BioLegend, RRID: AB_961390) in 0.5%Saponin (Sigma-Aldrich) in PBA/E at 4°C for 20 min. Samples were analyzed on a FACS-Canto-Plus (BD) and data were analyzed with FlowJo V10.7 (BD, Ashland, Oregon, USA). S-Protein-specific T_H_ cells are depicted as CD137^+^CD154^+^ among living CD4^+^CD3^+^. Representative FACS plots are shown in (*Suppl.Fig. 2*).

### Multi-dimensional flow cytometry analyses

The Uniform Manifold Approximation and Projection (UMAP) algorithm and the FlowSOM algorithm were used for unsupervised high dimensional flow cytometric analyses of the entire dataset with FlowJo version 10.8.1. Proportionally down-sampled single cells/ live/ CD3^+^/ CD4^+^ populations for each FCS file were concatenated in one single FCS file. UMAP was used for dimensionality reduction by using Euclidean as distance function with 15 Nearest Neighbors and 0.5 minimum distance. The following markers were used for building tSNE maps: 4-1BB, CD154, TNF, IFNγ, IL-4, and IL-17a. Resulting UMAP maps were fed into the FlowSOM (37). To identify clusters, heatmaps were built with median fluorescence intensity (MFI) values from each marker.

### Statistical analysis

All statistical analyses were performed in the analysis population as indicated and stratified by age, PCR-status, symptomatic disease, sex or sero-status from the serological assays if applicable. Descriptive analyses included the calculation of mean with standard deviation (SD) and medians with minimum and maximum or interquartile range (IQR) values for continuous variables, and absolute counts (n, with percentages) for categorical variables. Owing to the great data completeness, we performed no data imputations. All reported p-values are unadjusted and two-sided. Time course experiments were analyzed with the Friedman test and Dunn’s post hoc test for multiple comparisons. Antibody ratios were compared with the students t-test with Welch correction. Non-parametric estimation of Spearman’s rank correlation was performed with the following strength cut-offs ≥ 0.8 as very strong; ≥0.6-0.8 as moderately strong; ≥0.3-0.6 as fair and < 0.3 as poor (adapted from (65)). For comparison of two paired samples, the Wilcoxon-matched-pairs signed rank test as implemented in GraphPAD PRISM 9 was used. When more than two paired groups were compared, non-parametric Kruskal-Wallis with the Dunns post-hoc test was used.

### Study approval

The study was conducted according to the current version of the Declaration of Helsinki and has been approved by the institutional ethics committees of the Jena University Hospital and the respective data protection commissioner (approval number 2020-1776) and the ethics committee of the Thuringian chamber of physicians. All data were collected with unique pseudonyms on paper case report forms. These identifiers were later used to merge the questionnaire information with the laboratory information in an electronic study database. The study is registered at the German Clinical Trials Register: DRKS00022416.

## Data Availability

All data produced in the present work are contained in the manuscript

## AUTHOR CONTRIBUTIONS

CS, NA, MWP and SW had full access to all of the data in the study and take responsibility for the integrity of the data and the accuracy of the data analysis.

*Study concept and design*: MWP, SW, TK

*Acquisition of data*: MB, CS, SG, JG, MK, HP, SW, MWP

*Analysis and interpretation of data*: All authors.

*Drafting of the manuscript*: CS, NA, SW

*Critical manuscript revision & additional important intellectual content, data interpretation*: all authors.

*Statistical Analyses*: CS, SW

*Obtained funding*: MWP, SW

*Administrative, technical, or material support*: TK, SK, BL

*Study supervision*: MWP, SW

## ACKNOWLEDGEMENT

<u>CoNAN Study Group:</u> The project was carried out in cooperation with the district administration and the health department of the Ilm district.

Technische Universität Ilmenau, Ilmenau, Germany: Thomas Hotz;

Local Cooperation partners: Petra Enders, Renate Koch, Steffen Mai, Matthias Ullrich, Dagmar Rimek.

Institute of Clinical Chemistry and Laboratory Diagnostics and Integrated Biobank Jena (IBBJ), Jena University Hospital—Friedrich Schiller University, Jena, Germany: Cora Richert, Cornelius Eibner, Bettina Meinung, Kay Stötzer, Julia Köhler.

Children’s Hospital, Jena University Hospital—Friedrich Schiller University, Jena, Germany: Hans Proquitté, Hans Cipowicz, Christine Pinkwart.

Department of Anesthesiology and Intensive Care Medicine Jena University Hospital— Friedrich Schiller University, Jena, Germany: Michael Bauer, Petra Dickmann, Annika Licht, Juliane Scholz, Wibke Wetzker.

Institute for Infectious Disease and Infection Control, Jena University Hospital—Friedrich Schiller University, Jena, Germany: Gabi Hanf, Jasmin Müller, Jennifer Kosenkow, Franziska Röstel, Juliane Ankert, Aurelia Kimmig. Stefan Hagel. Christina Forstner.

Institute of Immunology, Jena University Hospital—Friedrich Schiller University, Jena, Germany: Raphaela Marquardt.

Institute of Medical Microbiology, Jena University Hospital—Friedrich Schiller University, Jena, Germany: Stefanie Deinhardt-Emmer, Sebastian Kuhn.

## REFERENCES

1. Osuchowski MF, Winkler MS, Skirecki T, Cajander S, Shankar-Hari M, Lachmann G, et al. The COVID-19 puzzle: deciphering pathophysiology and phenotypes of a new disease entity. Lancet Respir Med. 2021;9(6):622–42.

2. Cromer D, Juno JA, Khoury D, Reynaldi A, Wheatley AK, Kent SJ, et al. Prospects for durable immune control of SARS-CoV-2 and prevention of reinfection. Nat Rev Immunol. 2021;21(6):395–404.

3. Dan JM, Mateus J, Kato Y, Hastie KM, Yu ED, Faliti CE, et al. Immunological memory to SARS-CoV-2 assessed for up to 8 months after infection. Science. 2021;371(6529).

4. Seow J, Graham C, Merrick B, Acors S, Pickering S, Steel KJA, et al. Longitudinal observation and decline of neutralizing antibody responses in the three months following SARS-CoV-2 infection in humans. Nat Microbiol. 2020;5(12):1598–607.

5. Zhou F, Yu T, Du R, Fan G, Liu Y, Liu Z, et al. Clinical course and risk factors for mortality of adult inpatients with COVID-19 in Wuhan, China: a retrospective cohort study. Lancet. 2020;395(10229):1054–62.

6. Richardson S, Hirsch JS, Narasimhan M, Crawford JM, McGinn T, Davidson KW, et al. Presenting Characteristics, Comorbidities, and Outcomes Among 5700 Patients Hospitalized With COVID-19 in the New York City Area. JAMA 2020;323(20):2052–9.

7. Chansaenroj J, Yorsaeng R, Puenpa J, Wanlapakorn N, Chirathaworn C, Sudhinaraset N, et al. Long-term persistence of severe acute respiratory syndrome coronavirus 2 (SARS-CoV-2) spike protein-specific and neutralizing antibodies in recovered COVID-19 patients. PLoS One. 2022;17(4):e0267102.

8. Weis S, Scherag A, Baier M, Kiehntopf M, Kamradt T, Kolanos S, et al. Antibody response using six different serological assays in a completely PCR-tested community after a coronavirus disease 2019 outbreak-the CoNAN study. Clin Microbiol Infect. 2021;27(3):470 e1–e9.

9. Robbiani DF, Gaebler C, Muecksch F, Lorenzi JCC, Wang Z, Cho A, et al. Convergent antibody responses to SARS-CoV-2 in convalescent individuals. Nature. 2020;584(7821):437–42.

10. Lou B, Li TD, Zheng SF, Su YY, Li ZY, Liu W, et al. Serology characteristics of SARS-CoV-2 infection after exposure and post-symptom onset. Eur Respir J. 2020;56(2).

11. Padoan A, Cosma C, Sciacovelli L, Faggian D, and Plebani M. Analytical performances of a chemiluminescence immunoassay for SARS-CoV-2 IgM/IgG and antibody kinetics. Clin Chem Lab Med. 2020;58(7):1081–8.

12. Glockner S, Hornung F, Baier M, Weis S, Pletz MW, Deinhardt-Emmer S, et al. Robust Neutralizing Antibody Levels Detected after Either SARS-CoV-2 Vaccination or One Year after Infection. Viruses. 2021;13(10).

13. Wajnberg A, Amanat F, Firpo A, Altman DR, Bailey MJ, Mansour M, et al. Robust neutralizing antibodies to SARS-CoV-2 infection persist for months. Science. 2020;370(6521):1227–30.

14. Almendro-Vázquez P, Laguna-Goya R, Ruiz-Ruigomez M, Utrero-Rico A, Lalueza A, Maestro de la Calle G, et al. Longitudinal dynamics of SARS-CoV-2-specific cellular and humoral immunity after natural infection or BNT162b2 vaccination. PLoS Pathog. 2021;17(12):e1010211.

15. Sherina N, Piralla A, Du L, Wan H, Kumagai-Braesch M, Andrell J, et al. Persistence of SARS-CoV-2-specific B and T cell responses in convalescent COVID-19 patients 6-8 months after the infection. Med (N Y). 2021;2(3):281–95 e4.

16. Rydyznski Moderbacher C, Ramirez SI, Dan JM, Grifoni A, Hastie KM, Weiskopf D, et al. Antigen-Specific Adaptive Immunity to SARS-CoV-2 in Acute COVID-19 and Associations with Age and Disease Severity. Cell. 2020;183(4):996–1012.e19.

17. Carsetti R, Zaffina S, Piano Mortari E, Terreri S, Corrente F, Capponi C, et al. Different Innate and Adaptive Immune Responses to SARS-CoV-2 Infection of Asymptomatic, Mild, and Severe Cases. Front Immunol. 2020;11:610300.

18. Wang F, Hou H, Luo Y, Tang G, Wu S, Huang M, et al. The laboratory tests and host immunity of COVID-19 patients with different severity of illness. JCI Insight. 2020;5(10).

19. Peluso MJ, Deitchman AN, Torres L, Iyer NS, Munter SE, Nixon CC, et al. Long-term SARS-CoV-2-specific immune and inflammatory responses in individuals recovering from COVID-19 with and without post-acute symptoms. Cell Rep 2021;36(6):109518.

20. Anft M, Paniskaki K, Blazquez-Navarro A, Doevelaar A, Seibert FS, Hoelzer B, et al. COVID-19 progression is potentially driven by T cell immunopathogenesis. medRxiv. 2020:2020.04.28.20083089.

21. Sattler A, Angermair S, Stockmann H, Heim KM, Khadzhynov D, Treskatsch S, et al. SARS-CoV-2-specific T cell responses and correlations with COVID-19 patient predisposition. J Clin Invest. 2020;130(12):6477–89.

22. Wu J, Liang B, Chen C, Wang H, Fang Y, Shen S, et al. SARS-CoV-2 infection induces sustained humoral immune responses in convalescent patients following symptomatic COVID-19. Nat Comm. 2021;12(1):1813.

23. Cohen KW, Linderman SL, Moodie Z, Czartoski J, Lai L, Mantus G, et al. Longitudinal analysis shows durable and broad immune memory after SARS-CoV-2 infection with persisting antibody responses and memory B and T cells. Cell Rep Med. 2021;2(7):100354.

24. Braun J, Loyal L, Frentsch M, Wendisch D, Georg P, Kurth F, et al. SARS-CoV-2-reactive T cells in healthy donors and patients with COVID-19. Nature. 2020;587(7833):270–4.

25. Luo H, Camilleri D, Garitaonandia I, Djumanov D, Chen T, Lorch U, et al. Kinetics of anti-SARS-CoV-2 IgG antibody levels and potential influential factors in subjects with COVID-19: A 11-month follow-up study. Diagn Microbiol Infect Dis. 2021;101(4):115537.

26. Mak WA, Koeleman JGM, van der Vliet M, Keuren F, and Ong DSY. SARS-CoV-2 antibody and T cell responses one year after COVID-19 and the booster effect of vaccination: A prospective cohort study. J Infect. 2022;84(2):171–8.

27. Zhao J, Yuan Q, Wang H, Liu W, Liao X, Su Y, et al. Antibody Responses to SARS-CoV-2 in Patients With Novel Coronavirus Disease 2019. Clin Infect Dis. 2020;71(16):2027–34.

28. Zhang X, Lu S, Li H, Wang Y, Lu Z, Liu Z, et al. Viral and Antibody Kinetics of COVID-19 Patients with Different Disease Severities in Acute and Convalescent Phases: A 6-Month Follow-Up Study. Virol Sin. 2020;35(6):820–9.

29. Choe PG, Kim KH, Kang CK, Suh HJ, Kang E, Lee SY, et al. Antibody Responses One Year after Mild SARS-CoV-2 Infection. J Korean Med Sci. 2021;36(21):e157.

30. Demaret J, Lefevre G, Vuotto F, Trauet J, Duhamel A, Labreuche J, et al. Severe SARS-CoV-2 patients develop a higher specific T-cell response. Clin Transl Immunol. 2020;9(12):e1217.

31. Venet F, Gossez M, Bidar F, Bodinier M, Coudereau R, Lukaszewicz AC, et al. T cell response against SARS-CoV-2 persists after one year in patients surviving severe COVID-19. EBioMedicine. 2022;78:103967.

32. Breton G, Mendoza P, Hagglof T, Oliveira TY, Schaefer-Babajew D, Gaebler C, et al. Persistent cellular immunity to SARS-CoV-2 infection. J Exp Med 2021;218(4).

33. Barouch DH, Stephenson KE, Sadoff J, Yu J, Chang A, Gebre M, et al. Durable Humoral and Cellular Immune Responses 8 Months after Ad26.COV2.S Vaccination. N Engl J Med. 2021;385(10):951–3.

34. Krutikov M, Palmer T, Tut G, Fuller C, Azmi B, Giddings R, et al. Prevalence and duration of detectable SARS-CoV-2 nucleocapsid antibodies in staff and residents of long-term care facilities over the first year of the pandemic (VIVALDI study): prospective cohort study in England. Lancet Healthy Longev. 2022;3(1):e13–e21.

35. Grandjean L, Saso A, Torres Ortiz A, Lam T, Hatcher J, Thistlethwayte R, et al. Long-Term Persistence of Spike Protein Antibody and Predictive Modeling of Antibody Dynamics After Infection With Severe Acute Respiratory Syndrome Coronavirus 2. Clin Infect Dis. 2022;74(7):1220–9.

36. Garcia-Abellan J, Padilla S, Fernandez-Gonzalez M, Garcia JA, Agullo V, Andreo M, et al. Antibody Response to SARS-CoV-2 is Associated with Long-term Clinical Outcome in Patients with COVID-19: a Longitudinal Study. J Clin Immunol. 2021;41(7):1490–501.

37. Van Gassen S, Callebaut B, Van Helden MJ, Lambrecht BN, Demeester P, Dhaene T, et al. FlowSOM: Using self-organizing maps for visualization and interpretation of cytometry data. Cytometry A. 2015;87(7):636–45.

38. Stepanek L, Janosikova M, Stepanek L, Nakladalova M, and Borikova A. The kinetics and predictors of anti-SARS-CoV-2 antibodies up to 8 months after symptomatic COVID-19: A Czech cross-sectional study. J Med Virol. 2022.

39. Lau EH, Hui DS, Tsang OT, Chan WH, Kwan MY, Chiu SS, et al. Long-term persistence of SARS-CoV-2 neutralizing antibody responses after infection and estimates of the duration of protection. EClin Med. 2021;41:101174.

40. Schiffner J, Backhaus I, Rimmele J, Schulz S, Mohlenkamp T, Klemens JM, et al. Long-Term Course of Humoral and Cellular Immune Responses in Outpatients After SARS-CoV-2 Infection. Front Public Health. 2021;9:732787.

41. Khoury DS, Cromer D, Reynaldi A, Schlub TE, Wheatley AK, Juno JA, et al. Neutralizing antibody levels are highly predictive of immune protection from symptomatic SARS-CoV-2 infection. Nat Med. 2021;27(7):1205–11.

42. Rank A, Tzortzini A, Kling E, Schmid C, Claus R, Loll E, et al. One Year after Mild COVID-19: The Majority of Patients Maintain Specific Immunity, But One in Four Still Suffer from Long-Term Symptoms. J Clin Med. 2021;10(15).

43. Gallais F, Gantner P, Bruel T, Velay A, Planas D, Wendling MJ, et al. Evolution of antibody responses up to 13 months after SARS-CoV-2 infection and risk of reinfection. EBioMedicine. 2021;71:103561.

44. Murchu EO, Byrne P, Walsh KA, Carty PG, Connolly M, De Gascun C, et al. Immune response following infection with SARS-CoV-2 and other coronaviruses: A rapid review. Rev Med Virol. 2021;31(2):e2162.

45. Zhao J, Alshukairi AN, Baharoon SA, Ahmed WA, Bokhari AA, Nehdi AM, et al. Recovery from the Middle East respiratory syndrome is associated with antibody and T-cell responses. Sci Immunol. 2017;2(14).

46. Alshukairi AN, Khalid I, Ahmed WA, Dada AM, Bayumi DT, Malic LS, et al. Antibody Response and Disease Severity in Healthcare Worker MERS Survivors. Emerg Infect Dis. 2016;22(6).

47. Hamady A, Lee J, and Loboda ZA. Waning antibody responses in COVID-19: what can we learn from the analysis of other coronaviruses? Infection. 2022;50(1):11–25.

48. Tang F, Quan Y, Xin ZT, Wrammert J, Ma MJ, Lv H, et al. Lack of peripheral memory B cell responses in recovered patients with severe acute respiratory syndrome: a six-year follow-up study. J Immunol. 2011;186(12):7264–8.

49. Anderson DE, Tan CW, Chia WN, Young BE, Linster M, Low JH, et al. Lack of cross-neutralization by SARS patient sera towards SARS-CoV-2. Emerg Microbes Infect. 2020;9(1):900–2.

50. Sekine T, Perez-Potti A, Rivera-Ballesteros O, Stralin K, Gorin JB, Olsson A, et al. Robust T Cell Immunity in Convalescent Individuals with Asymptomatic or Mild COVID-19. Cell. 2020;183(1):158–68 e14.

51. Bilich T, Nelde A, Heitmann JS, Maringer Y, Roerden M, Bauer J, et al. T cell and antibody kinetics delineate SARS-CoV-2 peptides mediating long-term immune responses in COVID-19 convalescent individuals. Sci Transl Med. 2021;13(590).

52. Peng Y, Mentzer AJ, Liu G, Yao X, Yin Z, Dong D, et al. Broad and strong memory CD4(+) and CD8(+) T cells induced by SARS-CoV-2 in UK convalescent individuals following COVID-19. Nat Immunol. 2020;21(11):1336–45.

53. Grifoni A, Sidney J, Vita R, Peters B, Crotty S, Weiskopf D, et al. SARS-CoV-2 human T cell epitopes: Adaptive immune response against COVID-19. Cell host & microbe. 2021;29(7):1076–92.

54. Tarke A, Sidney J, Kidd CK, Dan JM, Ramirez SI, Yu ED, et al. Comprehensive analysis of T cell immunodominance and immunoprevalence of SARS-CoV-2 epitopes in COVID-19 cases. Cell Rep Med. 2021;2(2):100204.

55. Boppana S, Qin K, Files JK, Russell RM, Stoltz R, Bibollet-Ruche F, et al. SARS-CoV-2-specific circulating T follicular helper cells correlate with neutralizing antibodies and increase during early convalescence. PLoS Pathog. 2021;17(7):e1009761.

56. Stephenson E, Reynolds G, Botting RA, Calero-Nieto FJ, Morgan MD, Tuong ZK, et al. Single-cell multi-omics analysis of the immune response in COVID-19. Nat Med. 2021;27(5):904–16.

57. Juno JA, Tan HX, Lee WS, Reynaldi A, Kelly HG, Wragg K, et al. Humoral and circulating follicular helper T cell responses in recovered patients with COVID-19. Nat Med. 2020;26(9):1428–34.

58. Jung JH, Rha MS, Sa M, Choi HK, Jeon JH, Seok H, et al. SARS-CoV-2-specific T cell memory is sustained in COVID-19 convalescent patients for 10 months with successful development of stem cell-like memory T cells. Nat Comm. 2021;12(1):4043.

59. Loyal L, Braun J, Henze L, Kruse B, Dingeldey M, Reimer U, et al. Cross-reactive CD4(+) T cells enhance SARS-CoV-2 immune responses upon infection and vaccination. Science. 2021;374(6564):eabh1823.

60. Meyer-Arndt L, Schwarz T, Loyal L, Henze L, Kruse B, Dingeldey M, et al. Cutting Edge: Serum but Not Mucosal Antibody Responses Are Associated with Pre-Existing SARS-CoV-2 Spike Cross-Reactive CD4(+) T Cells following BNT162b2 Vaccination in the Elderly. J Immunol. 2022;208(5):1001–5.

61. Wirsching S, Harder L, Heymanns M, Grondahl B, Hilbert K, Kowalzik F, et al. Long-Term, CD4(+) Memory T Cell Response to SARS-CoV-2. Front Immunol. 2022;13:800070.

62. Yang HS, Costa V, Racine-Brzostek SE, Acker KP, Yee J, Chen Z, et al. Association of Age With SARS-CoV-2 Antibody Response. JAMA Netw Open. 2021;4(3):e214302.

63. Zeng F, Wu M, Wang J, Li J, Hu G, and Wang L. Over 1-year duration and age difference of SARS-CoV-2 antibodies in convalescent COVID-19 patients. J Med Virol. 2021;93(12):6506–11.

64. Callow KA, Parry HF, Sergeant M, and Tyrrell DA. The time course of the immune response to experimental coronavirus infection of man. Epidemiol Infect. 1990;105(2):435–46.

65. Chan YH. Biostatistics 104: correlational analysis. Singapore Med J. 2003;44(12):614–9.

