## Supplement for "PERSISTENT IMMUNITY AFTER MILD SARS CoV-2 INFECTION - THE CoNAN-LONG TERM STUDY -"

### SUPPLEMENTAL MATERIAL

Clara Schnizer[1*], Nico Andreas[2*], Wolfgang Vivas[1,3], Thomas Kamradt[2], Michael Baier[4], Michael Kiehntopf[5], Stefan Glöckner[4], André Scherag[6], Bettina Löffler[4], Steffi Kolanos[1], Joel Guerra[1,7] Mathias W. Pletz[1,8#], Sebastian Weis[1,3,7#] and the CoNAN study group.

[*#] contributed equally

[1] Institute for Infectious Diseases and Infection Control, Jena University Hospital- Friedrich Schiller University, Jena, Germany

[2] Institute of Immunology, Jena University Hospital- Friedrich Schiller University, Jena, Germany

[3] Leibniz Institute for Natural Product Research and Infection Biology – Hans Knöll Institute (HKI), Jena, Germany

[4] Institute of Medical Microbiology, Jena University Hospital- Friedrich Schiller University, Jena, Germany

[5] Institute of Clinical Chemistry and Laboratory Diagnostics and Integrated Biobank Jena (IBBJ), Jena University Hospital- Friedrich Schiller University, Jena, Germany

[6] Institute of Medical Statistics, Computer and Data Sciences, Jena University Hospital- Friedrich Schiller University, Jena, Germany

[7] Department of Anesthesiology and Intensive Care, Jena University Hospital- Friedrich Schiller University, Jena, Germany

[8] Center for Sepsis Control and Care, Jena University Hospital- Friedrich Schiller University, Jena, Germany

#### SUPPLEMENTAL MATERIAL

**Supplementary Figure 1** (refers to *Figure 2*): **A**) Ratios of anti-SARS-CoV-2 antibody levels over time from the indicated tests. **B,C**) ofaAnti-SARS-CoV-2 antibody levels over time as assessed with **B**) the quantitative Diasorin and **C**) the Euroimmun test (OD-ratio) and stratified by age, symptomatic disease, PCR status during the outbreak and sex. Dots represent individual participants. N= number of individuals per group. Friedman test with Dunns post-hoc analysis. * p<0.05; ** p<0.01; *** p<0.001; **** p<0.0001.

<u>Abbreviations</u>: ns..non-significant (p≥0.05).

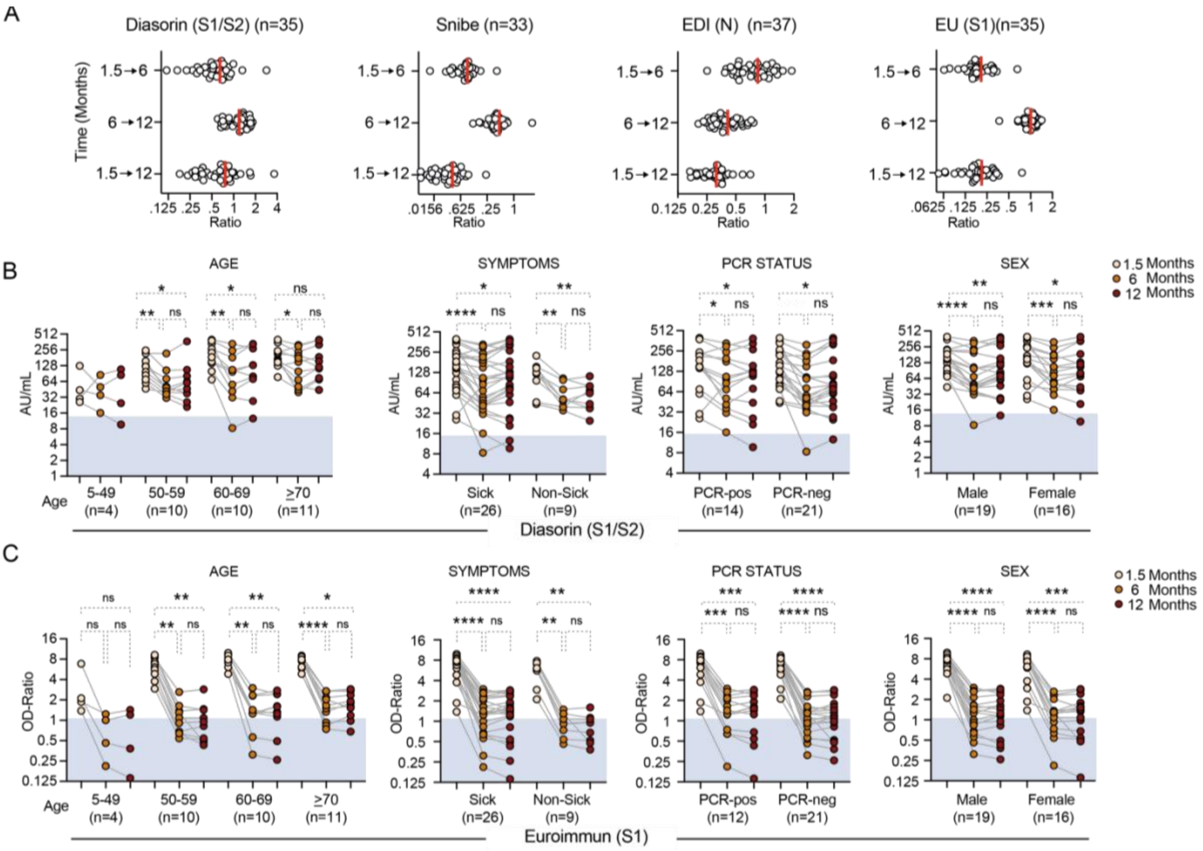

**Supplementary Figure 2** (refers to *Figure 3*): **A**) Representative gating on spike peptide (S.Pep1 (N) restimulated PBMCs. **B**) Gating stragegy for CD137 and CD154 in $CD3^{+}CD4^{+}$ cells in SARS-CoV-2 positive cells defined as previously PCR-positive or at least two positive antibody tests 1.5 months after SARS-CoV-2 outbreak and controls defined as no positive PCR results and less then two positive antibody tests after 1.5 and 12 months after after SARS-CoV-2 outbreak, respectively.

Abbreviations: DMSO.. Dimethylsulfoxide

A

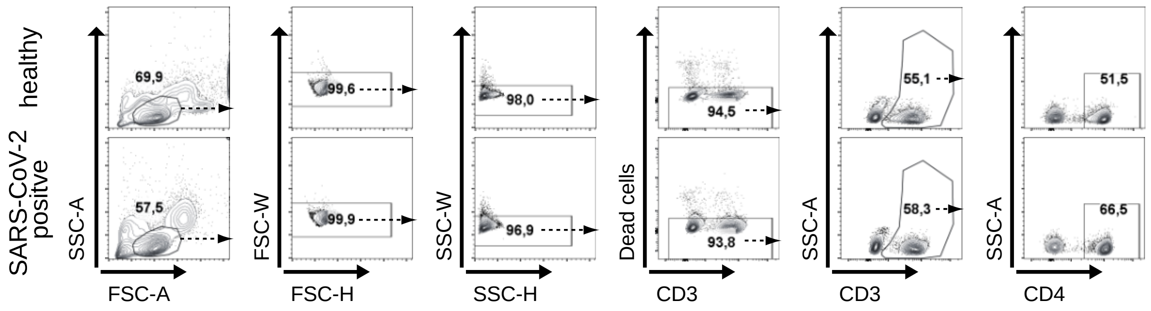

B

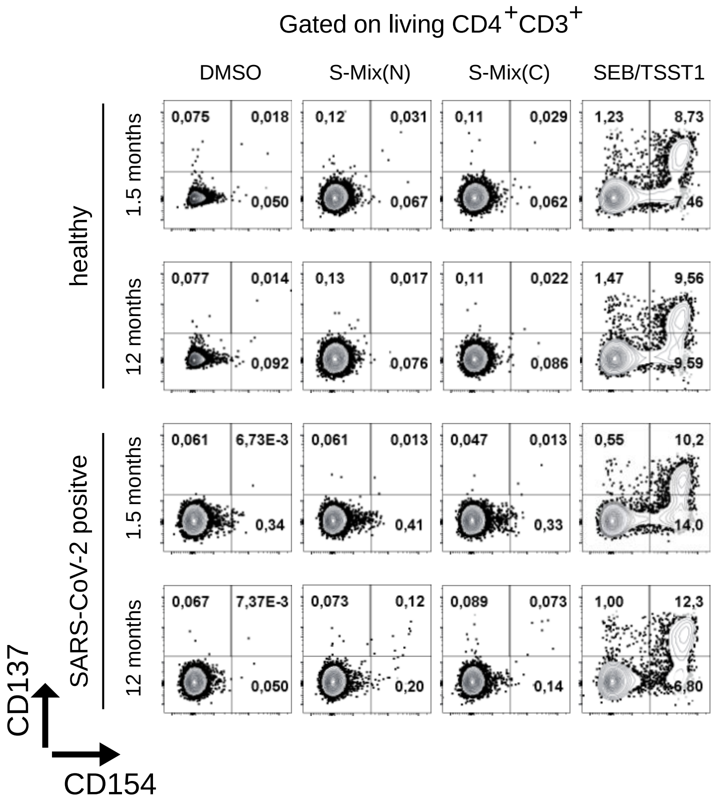

**Supplementary Figure 3** (refers to *Figure 3*): SARS-CoV-2 specific $T_H$ cell response of participants that were (**A-D**) seropositive but had no positive PCR test or (**E-F**) were tested positive either by PCR or by serum antibodies. **A**) % of 154+4-1BB+ $T_H$ cells stimulated with DMSO or S.Pep.2 (C). **B**) Direct comparison of % of CD154+4-1BB+ $T_H$ cells stimulated with or S.Pep.2 (C). **C, D**) % of 154+4-1BB+ $T_H$ cells stimulated with **C**) S.Pep.1 (N) or **D**) S.Pep.2 (C) in SARS-CoV-2 negative (Ctrl) and positive (SARS-CoV-2+) participants. **E, F**) % of 154+4-1BB+ $T_H$ cells in PCR-positive antibody negative vs. PCR-negative antibody-positive participants stimulated with **E**) S.Pep.1 (N) or **F**) S.Pep.2 (C) in SARS-CoV-2 negative (Ctrl) and positive (SARS-CoV-2+) participants. * p<0.05; ** p<0.01; *** p<0.001; **** p<0.0001.

Abbreviations: AB.. antibody; Ctrl.. control; DMSO.. Dimethylsulfoxide;

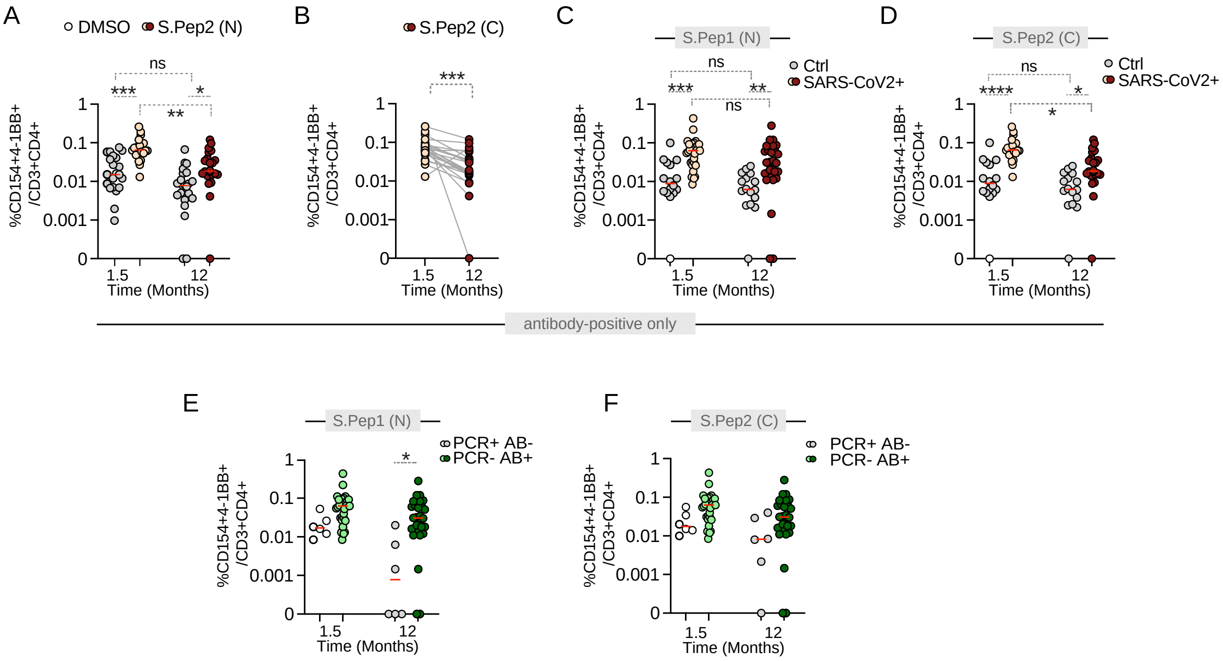

**Supplementary Figure 4** (refers to *Figure 4*): Single populations of indicated markers.

Abbreviations: CD.. cluster of differentiation; IL.. interleukin; INFγ.. interferon; TNF.. tumor necrosis factor.

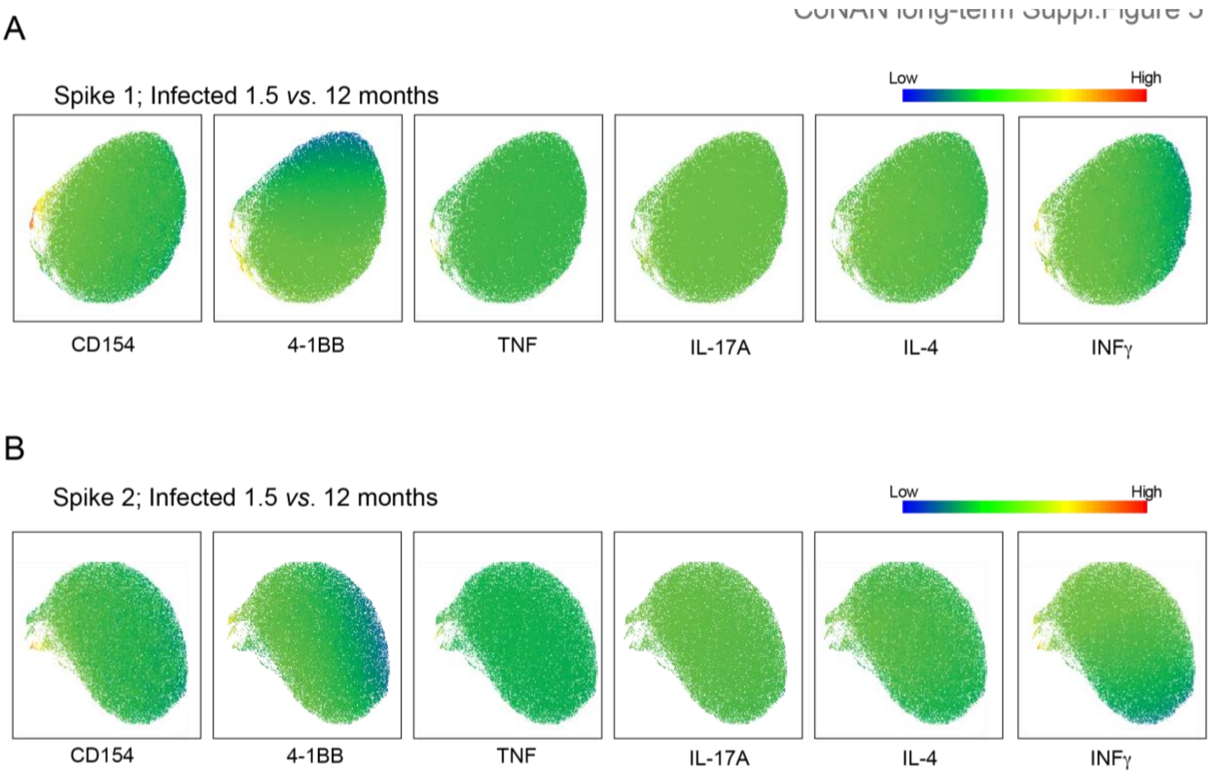

**Supplementary Figure 5** (refers to *Figure 5*): Spearman Correlation analysis of SARS-CoV-2 specific $T_H$ cells ($CD3^+CD4^+CD154^+4\text{-}1BB^+$) after S.Pep-1 (N) stimulation with all serological test results regardless of positive or negative antibody test results as indicated. Comparisons were the different combinations of the 1.5-month and 12-month time points after the SARS-CoV-2 outbreak in Neustadt am Rennsteig. Orange..1.5-month antibody data; Dark red.. 12-month antibody data.

<u>Abbreviations</u>: mo..month; TH.. T helper cells. Dots indicate individual participants.

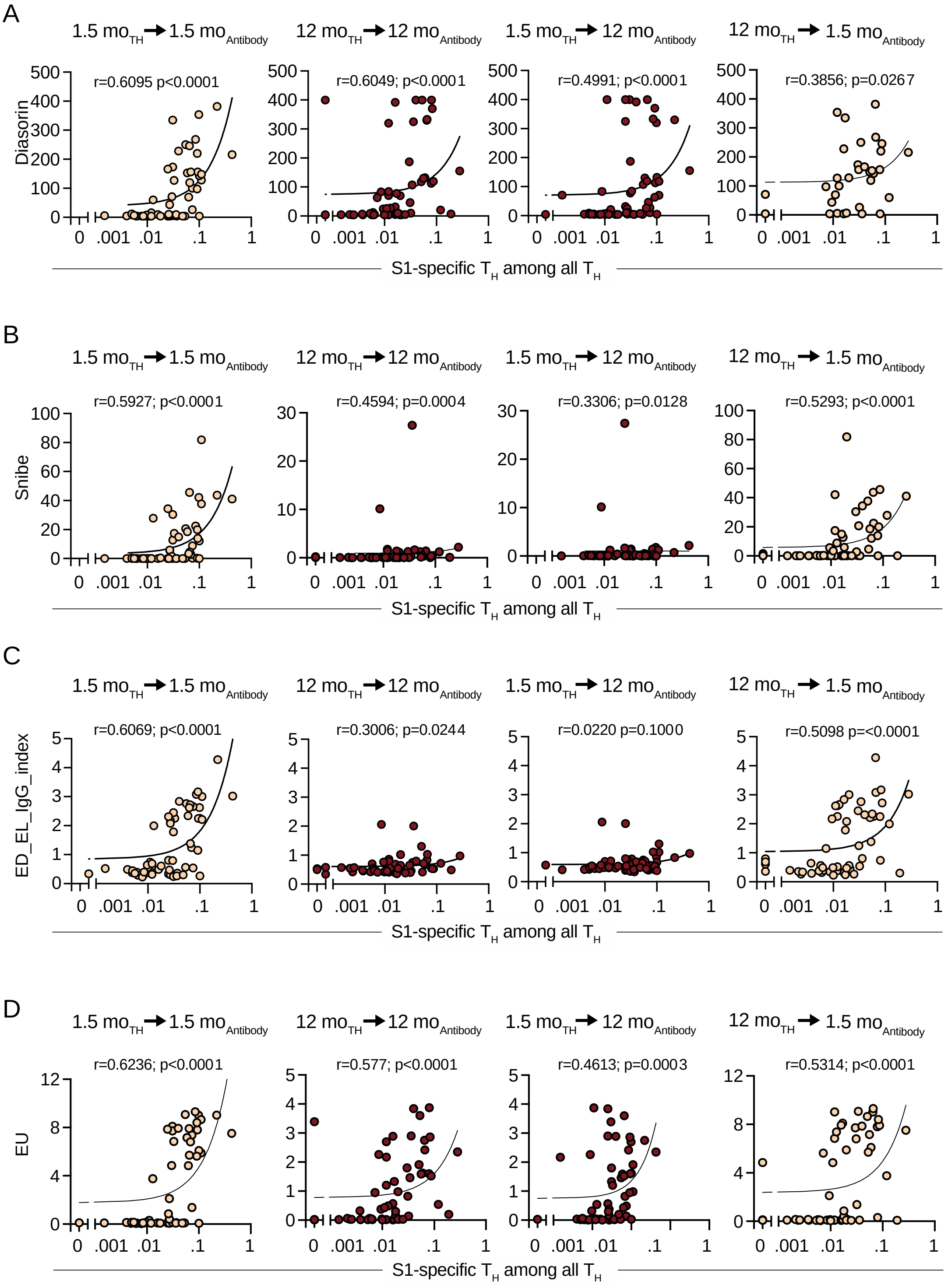

**<u>Supplementary Table 1:</u>** Sensitivities and specificities of the applied tests as provided by the manufacturer.

| Test and manufacturer | Instrument and manufacturer | Target | Sensitivity | Specificity | Note |
|---|---|---|---|---|---|
| **Enzyme-linked immunosorbent assays** | | | | | |
| EDI Novel Coronavirus COVID-19 IgG ELISA *Epitope Diagnostics, San Diego, USA* | Immunomat *Virion\Serion, Würzburg, Germany* | Nucleocapsid | 98·4% (184/187) 95% CI: 95·4%; 99·5% | 99·8% (623/624) 95% CI: 99·1%; 99·97% | no information about time of positivity has been given |
| Euroimmun Anti-SARS-CoV-2-ELISA (IgG) *Euroimmun, Lübeck, Germany* | Tecan Sunrise *Tecan, Männedorf, Switzerland* | S1 domain | 90·0% (27/30) 95% CI: 74·4%; 96·5% | 100% (80/80) 95% CI: 95·4%; 100% | >21 days post-symptom onset |
| **Chemiluminescence assays** | | | | | |
| Liaison SARS-CoV-2 S1/S2 IgG CLIA *DiaSorin, Saluggia, Italy* | DiaSorin Liaison XL *DiaSorin, Saluggia, Italy* | S1/S2 domains | 97·6% (40/41) 95% CI: 87·4%; 99.6% | 99·3% (1082/1090) 95% CI: 98·6%; 99·6% | >14 days post-symptom onset |
| Maglumi 2019-nCoV IgG CLIA *Snibe, Shenzhen, China* | MAGLUMI 800 *Snibe, Shenzhen, China* | 2019-nCoV recombinant antigen S+N | 91·21% 95% CI: NA | 100% 95% CI: NA | no information about time of positivity has been given |
| Elecsys Anti-SARS-CoV-2 ECLIA *Roche, Basel, Switzerland* | Cobas 8000 *Roche, Basel, Switzerland* | Nucleocapsid | 100% (29/29) 95% CI: 88·3%; 100% | 99·8% (5262/5272) 95% CI: 99·7%; 99·9% | ≥14 days post-symptom onset |

**Supplementary Table 2:** (refers to *Figure 5*): Spearman Correlation analysis of SARS-CoV-2 specific $T_H$ cells (CD3+CD4+CD154+4-1BB+) after S.Pep-2 (C) stimulation with positive serological test only. Comparisons were the different combinations of the 1.5-month and 12-month time points after the SARS-CoV-2 outbreak in Neustadt am Rennsteig. red.. fair statistically significant correlation

| Comparison | Spearman r [95%-Confidence Intervall] | p-value |
|---|---|---|
| **DiaSorin Liaison SARS-CoV-2 S1/S2 IgG CLIA** | | |
| 1.5 months (Antibody)→ 1.5 months ($T_H$) | 0.27 [-0.14 to 0.60] | 0.1770 |
| 12 months (Antibody)→ 12 months ($T_H$) | 0.27 [-0.04 to 0.63] | 0.1514 |
| 1.5 months (Antibody)→ 12 months ($T_H$) | 0.34 [-0.04 to 0.63] | 0.0677 |
| 12 months (Antibody)→ → 1.5 months ($T_H$) | -0.01 [-0.40 to 0.38] | 0.9475 |
| **Snibe Maglumi 2019-nCoV IgG** | | |
| 1.5 months (Antibody)→ 1.5 months ($T_H$) | 0.25 [-0.18 to 0.59] | 0.2384 |
| 12 months (Antibody)→ 12 months ($T_H$) | 0.10 [-0.52 to 0.65] | 0.7614 |
| 1.5 months (Antibody)→ 12 months ($T_H$) | -0.20 [-0.70 to 0.44] | 0.5431 |
| 12 months (Antibody)→ → 1.5 months ($T_H$) | 0.06 [-0.35 to 0.46] | 0.7683 |
| **EDI Novel Coronavirus COVID-19 IgG** | | |
| 1.5 months (Antibody)→ 1.5 months ($T_H$) | 0.53 [0.16 to 0.77] | 0.0064 |
| 12 months (Antibody)→ 12 months ($T_H$) | -0.87 [not available] | 0.6667 |
| 1.5 months (Antibody)→ 12 months ($T_H$) | -1.00 [not available] | 0.3333 |
| 12 months (Antibody)→ → 1.5 months ($T_H$) | -0.13 [-0.58 to 0.29] | 0.5389 |
| **Euroimmun Anti-SARS-CoV-2-ELISA IgG** | | |
| 1.5 months (Antibody)→ 1.5 months ($T_H$) | 0.25 [-0.16 to 0.59] | 0.2115 |
| 12 months (Antibody)→ 12 months ($T_H$) | 0.21 [-0.24 to 0.59] | 0.3442 |
| 1.5 months (Antibody)→ 12 months ($T_H$) | -0.02 [-0.45 to 0.41] | 0.9225 |
| 12 months (Antibody)→ → 1.5 months ($T_H$) | -0.12 [-0.49 to 0.30] | 0.5788 |
| **Roche Elecsys Anti-SARS-CoV-2 ECLIA** | | |
| 1.5 months (Antibody)→ 1.5 months ($T_H$) | -0.06 [-0.42 to 0.33] | 0.7722 |
| 12 months (Antibody)→ 12 months ($T_H$) | -0.12 [-0.47 to 0.27] | 0.5466 |
| 1.5 months (Antibody)→ 12 months ($T_H$) | 0.25 [-0.14 to 0.57] | 0.1953 |
| 12 months (Antibody)→ → 1.5 months ($T_H$) | 0.25 [-0.14 to 0.57] | 0.1953 |

**Supplementary Table 3:** (refers to *Figure 5*): Spearman Correlation analysis of SARS-CoV-2 specific $T_H$ cells after S.Pep-2 (C) stimulation with all serological test results regardless of positive or negative antibody test results as indicated. Comparisons were the different combinations of the 1.5-month and 12-month time points. blue.. moderately strong & statistically significant correlation, red.. fair & statistically significant correlation.

| | **Spearman r [95%-Confidence Intervall]** | **p-value** |
|---|---|---|
| **DiaSorin Liaison SARS-CoV-2 S1/S2 IgG CLIA** | | |
| 1.5 months (Antibody)→ 1.5 months ($T_H$) | 0.60 [0.40 to 0.75] | <0.0001 |
| 12 months (Antibody)→ 12 months ($T_H$) | 0.44 [0.19 to 0.63] | 0.0008 |
| 1.5 months (Antibody)→ 12 months ($T_H$) | 0.55 [0.32 to 0.71] | <0.0001 |
| 12 months (Antibody)→ → 1.5 months ($T_H$) | 0.16[-0.12 to 0.41] | 0.2396 |
| **Snibe Maglumi 2019-nCoV IgG** | | |
| 1.5 months (Antibody)→ 1.5 months ($T_H$) | 0.62 [0.42 to 0.76] | <0.0001 |
| 12 months (Antibody)→ 12 months ($T_H$) | 0.19 [0.09 to 0.44] | 0.1698 |
| 1.5 months (Antibody)→ 12 months ($T_H$) | 0.39 [0.14 to 0.60] | 0.0028 |
| 12 months (Antibody)→ → 1.5 months ($T_H$) | 0.22 [-0.05 to 0.46] | 0.1019 |
| **EDI Novel Coronavirus COVID-19 IgG** | | |
| 1.5 months (Antibody)→ 1.5 months ($T_H$) | 0.64 [0.45 to 0.78] | <0.0001 |
| 12 months (Antibody)→ 12 months ($T_H$) | 0.02 [-0.26 to 0.28] | 0.9130 |
| 1.5 months (Antibody)→ 12 months ($T_H$) | 0.25 [-0.02 to 0.49] | 0.0651 |
| 12 months (Antibody)→ → 1.5 months ($T_H$) | 0.26 [-0.01 to 0.50] | 0.0503 |
| **Euroimmun Anti-SARS-CoV-2-ELISA IgG** | | |
| 1.5 months (Antibody)→ 1.5 months ($T_H$) | 0.62 [0.43 to 0.77] | <0.0001 |
| 12 months (Antibody)→ 12 months ($T_H$) | 0.42 [0.17 to 0.62] | 0.0014 |
| 1.5 months (Antibody)→ 12 months ($T_H$) | 0.47 [0.23 to 0.66] | 0.0003 |
| 12 months (Antibody)→ → 1.5 months ($T_H$) | 0.27 [0.002 to 0.51] | 0.0421 |
| **Roche Elecsys Anti-SARS-CoV-2 ECLIA** | | |
| 1.5 months (Antibody)→ 1.5 months ($T_H$) | 0.61 [0.41 to 0.76] | <0.0001 |
| 12 months (Antibody)→ 12 months ($T_H$) | 0.20 [-0.08 to 0.45] | 0.1434 |
| 1.5 months (Antibody)→ 12 months ($T_H$) | 0.59 [0.38 to 0.74] | <0.0001 |
| 12 months (Antibody)→ → 1.5 months ($T_H$) | 0.24 [-0.04 to 0.48] | 0.0809 |
